# Individual-based and neighbourhood-based socio-economic factors relevant for contact behaviour and epidemic control

**DOI:** 10.1101/2025.03.24.25324502

**Authors:** Laura Di Domenico, Martina L. Reichmuth, Christian L. Althaus

## Abstract

Identifying sources of heterogeneity in contact patterns is key to inform disease transmission models. Recent works have investigated how individual-based socio-economic factors, besides age, affect contact behaviour, but neglected the individuals’ area of living. Here, we aimed at estimating contact matrices stratified by both individual-based and area-based socio-economic factors. We used social contact data from Switzerland collected in 2021, combined with a neighbourhood-based index of socio-economic position (SEP). First, we found a positive association between education level and number of contacts in the elderly, and, notably, a negative association between SEP level and number of contacts in adults. Second, despite lacking socio-economic information on the contacts, we developed a method to reconstruct contact matrices fully stratified by age, education level, and SEP, with varying assortativity levels. Third, integrating the matrices into a transmission model revealed heterogeneous disease burden, with higher attack rates in adults with higher education level living in low SEP areas and seniors with higher education level living in high SEP areas. Adults and young individuals living in high SEP areas were the main contributors to transmission. We found that the less assortative contacts are, the higher the chances of a targeted strategy to be successful, and the lower the control effort required to prevent disease spread. Our results shed light on contact behaviour in previously neglected socio-economic groups, enable model integration of socio-economic indicators, and provide insights to improve disease control.

## Introduction

The COVID-19 pandemic has highlighted the value of infectious disease modeling incorporating data on human behaviour, such as social contact patterns, to evaluate the effectiveness of control strategies and inform public health policies^1^. Several modeling works integrated age-stratified contact matrices^2–6^, estimated from pandemic survey data^7,8^. However, contact patterns can vary not only by age but also by other variables, e.g. socio-economic factors (income, education level^9–11^), vaccination status^12–15^, and vaccine hesitancy^16^, that can all affect individuals’ risk perception and adaptive behaviour.

Population response to COVID-19 measures varied across socio-economic groups. Job market structure, income and house crowding acted as constraints in reducing mobility, shaping adherence to public health measures^17–19^. Socio-economic factors also influenced testing rates and response to change in testing guidelines^20,21^. Moreover, studies found socio-economic factors associated with COVID-19 disease severity^20,22^ and with vaccine uptake, both for COVID-19^23–26^ and other diseases^27,28^, showing that individuals with lower socio-economic status are less likely to be vaccinated. These inequities call for the integration of socio-economic heterogeneity in epidemic models^29–33^.

Recent epidemic modeling works included mixing patterns stratified by age and socio-economic variables^9,34,35^. However, these studies have focused on individual-based features of the survey participant (i.e., socio-economic status, or SES), neglecting the environment in which the individual lives (i.e., socio-economic position, or SEP). In Switzerland, a SEP index has been defined at building level^36,37^, as an area-based measure reflecting the socio-economic situation of the neighbourhood, intended as a collection of the closest 50 households. Previous work uncovered the association of the SEP index with health-related outcomes such as human papillomavirus (HPV) vaccine uptake^38^, rate of testing, infection, hospitalization, and mortality related to COVID-19^20^. As the SEP refers to the neighbourhood, rather than to individual-based characteristics (e.g. education level and income), it represents an additional dimension of socio-economic heterogeneity. By combining individual-based and area-based factors, it becomes possible to investigate contact behaviour in intersectional socio-economic groups so far overlooked, e.g. individuals with low SES living in high SEP areas and vice versa.

To integrate socio-economic factors into epidemic models, one convenient framework is to introduce an age-stratified contact matrix that can be further stratified by additional dimensions, such as SES and SEP^34^. However, estimating such a matrix from survey data requires knowledge of

socio-economic information related to both the participants and their contacts. The latter information is often missing in traditional social contact surveys, where participants only report the age of their contacts. This hampers the construction of a fully stratified contact matrix, and also opens questions about whether contacts along social dimensions are assortative, i.e., to what extent individuals engage contacts with individuals belonging to the same socio-economic group. Quantifying the assortativity of contacts is key to understanding the impact of socio-economic heterogeneity on contacts, and therefore on disease spread and epidemic control.

In this study, we aimed to extend traditional age-dependent social contact matrices with two complementary (individual-based and area-based) socio-economic dimensions. We used social contact data collected in 2021 in Switzerland with SES information for the survey participants, combined with a neighbourhood-based SEP index. First, we characterized the dependence of contact activity on socio-economic factors, distinguishing between SES and SEP, to identify determinants influencing the number of contacts. Secondly, we built contact matrices accounting for both age and socio-economic factors, by stratifying for both the participant and the contacts. To deal with missing socio-economic information on the peers, we developed a method to reconstruct fully stratified contact matrices based on the observed partial data, by preserving conditions on reciprocity and aggregation.

This method allowed us to identify ranges of contact assortativity across socio-economic groups. Third, we integrated the generated contact matrices into a Susceptible-Infectious-Recovered (SIR) epidemic model, to identify the most relevant socio-economic groups in terms of disease burden and transmission. Finally, we tested the effectiveness of a targeted strategy aimed at reducing contacts or increasing vaccination uptake in a specific socio-economic group. Through the concept of the type-reproduction number, we identified which targeted control strategies would be effective, and we exposed the relation between contact assortativity and control effort.

## Methods

### Data

#### Social contact data

We used Swiss contact data collected during the COVID-19 pandemic through the European CoMix survey^7^, containing individual-based socio-economic information (education level, income) and spatial information (municipality) for survey participants. The data are described in detail in another study^39^ and a subset of variables are available on Zenodo^40^. To increase the sample size, we aggregated data from three contact surveys involving adults, carried out in January, June and December 2021, for a total of 3552 adult participants. We also considered three contact surveys for individuals younger than 18 years, filled out by their parents, for a total of 910 children.

All participants in the survey reported the following information: age, gender, region (urban or rural), country of birth, education level, household income, employment status, household size, COVID-19 vaccination status, municipality of residence. We divided the participants into four age groups, i.e., 0-14, 15-24, 25-64, and 65+ year olds. We refer to these age groups as children, young adults, adults, and seniors, respectively. We aggregated the education level into two categories: high education level, and middle-low education level. The former group corresponds to the tertiary level in the Swiss Education System^41^, which includes advanced vocational education or a university degree (Bachelor, Master or PhD); the latter group includes individuals with upper-secondary education or without any post-compulsory education. All children were classified as middle-low education. Further details about participant covariates and survey periods are contained in the Supplementary Information (Fig. S1).

Participants reported their contacts. A contact was defined as anyone who met the participant in person with whom at least a few words were exchanged or physical contact was made. The age of the contacts was available for 28,436 out of 35,072 contacts declared (81%). If a participant did not report information on the age of the contact, we imputed one of the four age groups (0-14, 15-24, 25-64, 65+ year olds), proportionally to the observed distribution of contacts’ age for participants of the same group. Some participants reported contacts with age brackets overlapping two of the age groups under consideration (e.g. contacts declared as 0-18 year olds). As above, we sampled from the reported age group with a weighting consistent with the age distribution of contacts for the participants’ own age group.

#### SEP index

We used a neighbourooh-based index of socio-economic position, so-called SEP, that has been defined for each residential building in Switzerland. The SEP index is a composite measure of the socio-economic level of the neighbourhood (defined as the 50 closest households) based on four domains, i.e., rent per square meter, education level, occupation and overcrowding. For details on the construction of the index, see ^36,37^.

We assigned a SEP level (low or high) to each participant based on the municipality of residence declared in the survey, with the following approach. First, we computed a weighted average SEP index at municipality level. We created an aggregated index weighted by the number of individuals living in each residential building in the municipality of interest, using household population data^42^ (see Supplementary Information and Fig. S2 for details). Then, we used the weighted average SEP by municipality to group participants as low SEP or high SEP based on their municipality of residence, using the median as threshold value.

#### Population data

We aimed at stratifying the Swiss population by age group (0-14, 15-24, 25-64, and 65+ year olds), education level (high or middle-low), and SEP level (high or low). As more detailed data were not available, we extrapolated an estimate combining data at district level by age group^43^ or by education level^44^, with national data by education level and age group^44^. Details are contained in the Supplementary Information and Fig. S3.

### Statistical analysis

#### Contact activity by group

We first computed the crude mean number of contacts per participant by education level (high or middle-low) and SEP (high or low), for each age group, after truncation at 50 contacts to remove the impact of outliers. We displayed the results in a heatmap to uncover the groups with higher or lower number of contacts. We stratified the analysis by age group (Fig. 2). Heatmaps further stratified by survey wave can be found in the Supplementary Information (Fig. S4). We built bootstrapped confidence intervals for the mean number of contacts out of 1000 random samples with replacement of the participants data.

#### Regression model

We used a generalized linear model, to estimate the effect of an individual-based socio-economic factor, i.e., education level, and area-based SEP level on the number of contacts. We assumed a Poisson distribution to account for count data. In the main analysis, the response variable for the regression model was the overall number of contacts per participant. In a sensitivity analysis, we analyzed contacts outside the household, and contacts at work (Fig. S5). We adjusted for age group, panel wave, gender, region, country of birth, household income, household size, employment status, vaccination status, and type of day (weekday or weekend). In a sensitivity analysis, we also adjusted for population density (Fig. S6) and stratified by survey wave (Fig. S7). We included an interaction term between age and education level, and between age and SEP level, to account for effects stratified by age group. We then computed the combined relative risk (RR) as *exp*(β +β’), where β is the estimated coefficient for education level only (or SEP level only), and β’ is the estimated coefficient for the corresponding interaction term with age (Fig. 2). In a sensitivity analysis, we also included an interaction term between age and household income (Fig. S8).

### Expanded contact matrix

#### Age-stratified contact matrix

We estimated an age-stratified contact matrix from survey data, adjusting for population demography and contact reciprocity^45^. We did not adjust for weekend effect, as the proportion of participants surveyed during weekdays or weekends was representative of the weekly pattern. We derived a contact matrix *M* whose elements *M* represent the average number of contacts that one individual in age group *i* engages with individuals in age group *j*, with *i, j* being either 0-14, 15-24, 25-64, and 65+ years old. We corrected for reciprocity by defining the matrix 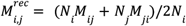, to ensure the symmetry 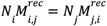, where N is the population size of age group *i*, and 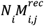 is the total number of contacts between age group *i* and age group *j*.

#### Expanded contact matrix

We aimed to estimate an expanded reciprocal contact matrix 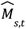 , with *s, t* being of the form (*i, v, d*), stratified by age group *i*, SEP level *v* (low or high) and education level *d* (middle-low or high). In the social contact survey, information on education level and SEP was available only for participants, while it was missing for contacts, for whom only age was available. We first built an intermediate contact matrix 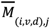 disaggregated on the participant side. This is a rectangular matrix informed from available data. The element 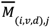 represents the average number of contacts of a participant in age group *i*, SEP level *v* and education level *d*, engaged with individuals belonging to age group *j*. We derived an adjusted contact matrix 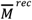 correcting by reciprocity, to be consistent with the reciprocal age-stratified contact matrix *M*^*rec*^after aggregating over socio-economic groups. Details on this reciprocity correction are presented in the Supplementary Information.

We further expanded the adjusted intermediate matrix 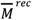 to a fully stratified contact matrix 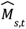. To derive the elements of the matrix 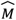 , we set a system of linear equations, with constraints due to the properties of the structure of the social contact matrix. In particular, the matrix 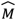 is required to fulfill conditions on (i) reciprocity, i.e., the total number of contacts 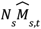 between individuals in group *s* and group *t* must be equal to 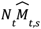 (ii) aggregation, i.e., the number of contacts summed over all socio-economic groups (excluding age) must be consistent with the elements of the adjusted intermediate contact matrix 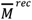 , so that 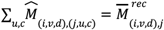 (iii) positivity, i.e., the number of contacts in each cell cannot be negative.

Practically, the global system can be solved through 10 independent linear systems corresponding to specific blocks of the contact matrix (see Fig. S9 for an illustration). In order to solve the system for each block, it is required to set the value of some parameters *q* ∈ (0, 1), that define the distribution of contacts across socio-economic groups (see Supplementary Information for the mathematical definition). They can be interpreted as assortativity parameters along SEP and education level dimension. Assortativity refers to the preferential behaviour of an individual in engaging more contacts with people with the same characteristics. Age-stratified contact matrices estimated from empirical data are generally assortative with age^45^, meaning that they have a stronger diagonal component with respect to homogeneous mixing, where the number of contacts is proportional to the size of the contact group and no preferential behaviour is in place.

We explored the parameter space through uniform random sampling in the range (0, 1) for each parameter *q*, and identified combinations of parameter values solving the system and leading to a positive expanded matrix. A set of combinations are displayed in Fig. S10-12. We generated 10,000 expanded contact matrices 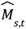 with varying assortativity parameters, that are all compatible with our partial data 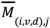. We then investigated the implications of these matrices in terms of population mixing and disease spreading.

#### Assortativity index

To characterize the mixing along the SEP and education level dimensions, we computed two assortativity indexes based on the 2×2 contact matrices 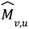 and 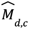 stratified by only SEP or only education level, respectively. These matrices are obtained from the expanded contact matrix 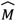 after aggregation. The aggregation is performed by summing along the rows and computing a weighted mean across the rows based on the group size. As assortativity index α, we decided to use the definition introduced in Ref.^46^ based on the trace of the matrix of proportions of contacts (see the Supplementary Information for the definition). The index α ∈ (0, 2) is defined so that: (i) α = 2 indicates fully assortative (or “segregate”) mixing, where individuals are in contact only with people in their same group, i.e., no contacts across different groups occur (corresponding to null matrix elements outside of the diagonal); (ii) α = 1 indicates proportional or homogeneous mixing^47^, i.e., the

per-capita contact rate 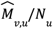 is constant for any group *u*, and therefore the number of contacts 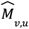 is proportional to the size of the contacted group N ; (iii) α = 0 represents fully disassortative mixing, i.e., no contacts within the same group occur (null matrix elements on the diagonal). We computed the assortativity index α for both 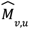 and 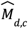 obtained from 10,000 different expanded contact matrices, and we compared it with an expanded contact matrix 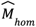 that assumes proportional mixing along SEP and education levels.

### Epidemic modeling

#### Compartmental model

We considered an extended version of an age-stratified Susceptible-Infectious-Recovered (*SIR*) compartmental model for disease transmission^48,49^. Assuming the same epidemiological parameters for all age groups, the force of infection on susceptible *S*_*i*_ of age group *i* can be written as 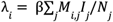 , where β is the transmission rate per contact, *M*_*i*,*j*_ is the age-stratified contact matrix introduced above, and *I_j_*/*N_*j*_* is the fraction of infectious individuals in age group *j*. Here, we extended the age stratification to a stratification by SEP and education level, where *S_*i,v,d*_* is the number of susceptible of age group *i*, SEP level *v*, education level *d*. Analogously, we integrated our estimated expanded contact matrix 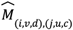 in the force of infection 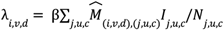.

#### Reproductive number and disease spread

The basic reproductive number *R*_0_ , defined as the average number of infections generated by a case in a fully susceptible population, is a key quantity which characterizes the epidemic dynamics^50^. For structured compartmental models, *R*_0_ can be computed as the dominant eigenvalue ρ(*K*) of the so-called next-generation matrix *K*^51,52^, which encodes the contact matrix *M* and epidemiological characteristics of the groups. When all groups are epidemiologically equivalent (i.e., having the same susceptibility, infectiousness, and recovery rate), the ratio ρ(*K*’)/ρ(*K*) is equal to ρ(*M*’)/ρ(*M*). Hence, given a couple of contact matrices *M* and *M*’, the relative variation in their dominant eigenvalue would correspond to the relative variation in *R*_0_ when integrating these matrices into the *SIR* model. We computed the relative variation in *R*_0_ for a set of expanded contact matrices 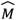, with respect to a matrix 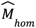 with proportional mixing in the SEP and education level dimension (i.e., number of contacts is proportional to the size of the contact group). We also computed the relative variation in *R*_0_ when adding one dimension to age-stratification, either SEP or education level only, to quantify their relative role. We expect this relative variation to be always positive (see^34^ and Supplementary information). For each group *s* = (*i, v, d*), we also computed its proportional contribution to *R*_0_ . In particular, for each group *s* we computed the cumulative elasticity *e*_*s*_ , following the framework described in Ref.^53^. The elasticity index allowed us to identify which groups contribute the most to transmission. Finally, we simulated the epidemic spread in absence of control measures and computed the relative attack rate in each group (i.e., the cumulative number of infections at the end of the epidemic), to identify the ones mostly affected by disease burden.

#### Type-reproduction number and targeted control strategies

In a population with homogenous mixing, the basic reproductive number is linked with the herd immunity threshold 1 − 1/*R*_0_, which represents the fraction of susceptible required to be fully immune to halt the epidemic^54^. For heterogeneous mixing, the type-reproduction number *T*_*g*_ has been introduced as a measure of the control effort needed to contain the epidemic when targeting one specific group *g* of the population^55,56^. The epidemic is controlled if a proportion of group *g* greater than 1 − 1/*T*_*g*_ is permanently immune or fully isolated at the start of the epidemic. Thus, the quantity 1 − 1/*T*_*g*_ corresponds to the immunity threshold specific to some target group(s). The type-reproduction number can be computed from the next-generation matrix (see Refs.^55,56^ and Supplementary Information for additional details).

We computed the type-reproduction number and the corresponding immunity threshold for a set of target groups, to test the effectiveness of targeted strategies. For the strategies allowing effective control of the epidemic, we computed the overall control effort, i.e., the fraction of targeted individuals over the whole population, to allow comparisons across strategies targeted to groups of different sizes.

## Results

### Contact activity by socio-economic group

For our analysis, we divided each age group in four socio-economic groups, i.e., individuals with high or middle-low education living in areas with high or low SEP. The size of each group in the general Swiss population is displayed in Fig. 1a. Higher densely populated areas typically have a higher average SEP compared to lower densely populated areas (Fig. 1a). The sample sizes of participants in the survey were found to be well representative of the general population (Fig. S2).

**Figure 1.**
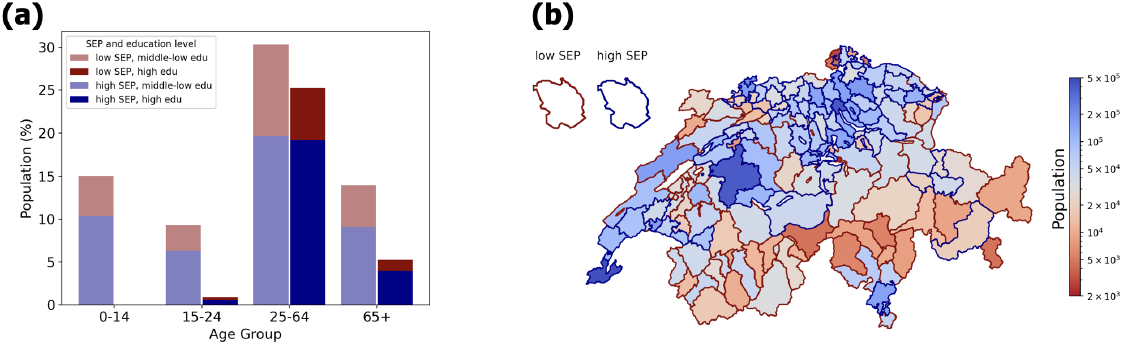
Swiss population. **(a)** Population size (in %) in Switzerland by age group (x-axis), SEP level (red/blue color), and education level (lighter/darker shade). The group of children (0-14 year olds) with high education level is empty. **(b)** Map of population size in Switzerland at district level. The face color of each district indicates the population; shades of blue and red indicate districts with larger or smaller population sizes, respectively. The edge color of each district indicates the SEP classification (either high SEP and low SEP, in blue and red respectively).

**Figure 2.**
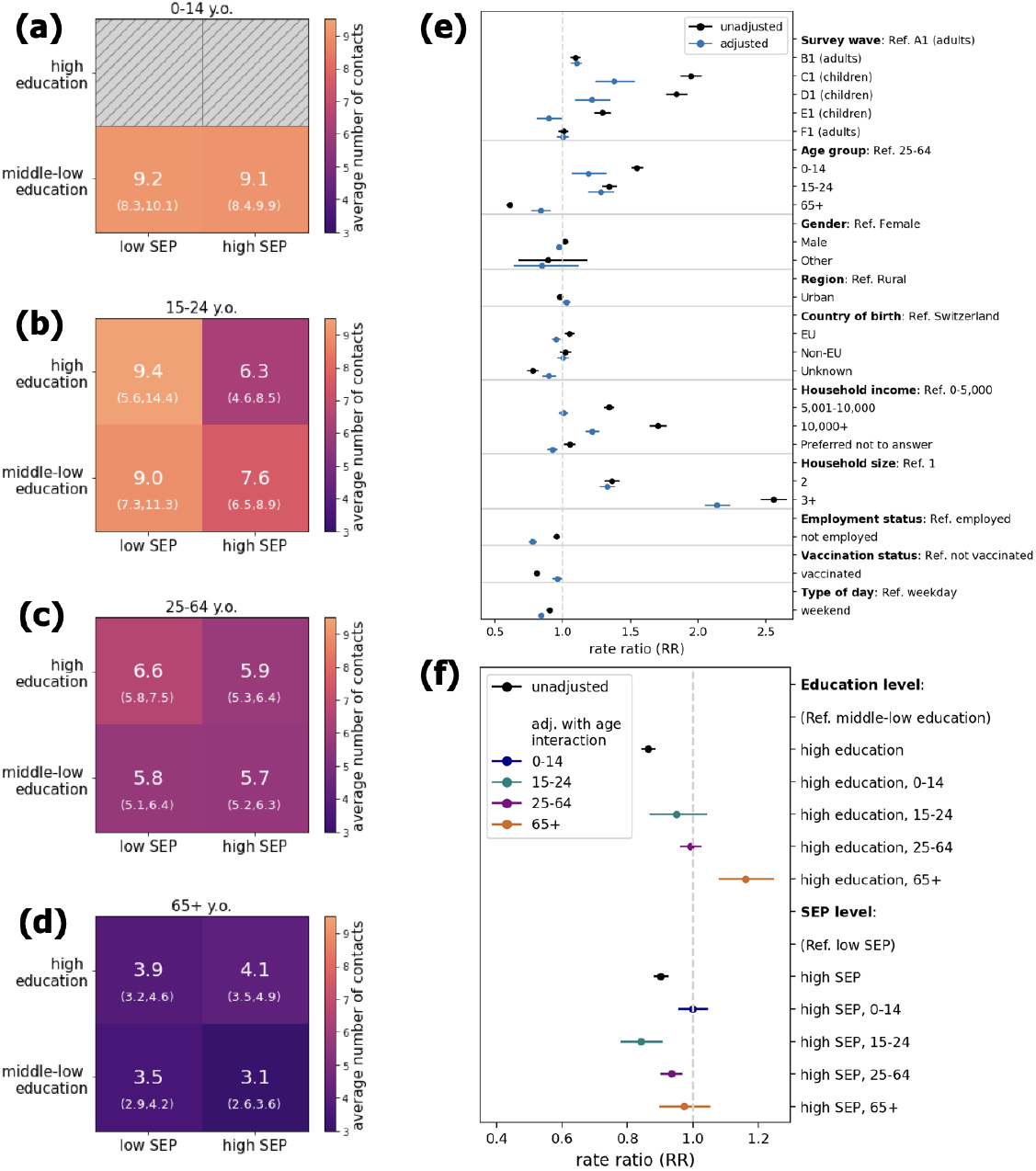
**(a-d)** Average number of contacts for each individual-based (education level) and area-based (SEP) socio-economic group. The heatmap shows the crude mean number of contacts (along with 95% confidence interval obtained with 1000 bootstraps) engaged by a participant belonging to one of the four socio-economic groups, depending on the age group **(a)** 0-14 years old (children), **(b)** 15-24 years old (young adults) **(c)** 25-64 years old (adults) and **(d)** 65+ years old (seniors). **(e-f)** Results of the regression analysis. The rate ratio (RR) represents the relative change in the outcome, i.e., the average number of contacts of a participant in a given group with respect to a group of reference. Black and colored dots represent the estimate obtained with a univariate and multivariate model respectively. Bars indicate 95% confidence intervals.

We computed the mean number of contacts per participant for each socio-economic group, distinguishing by age (Fig. 2a-d). Young adults (15-24 year olds) living in low SEP areas had more contacts with respect to those with high SEP. Among adults (25-64 year olds), the group with the highest contact activity were individuals with high education level living in low SEP areas. This was not observed for seniors (65+ year olds), where individuals with high education level engaged a similar number of contacts regardless of their SEP. The different contact patterns observed within each age group suggested a possible interaction effect between age and socio-economic factors in shaping contact activity. We included an interaction term with age for SEP and education level in a multivariate regression model, adjusting for household income, household size, employment status and other relevant variables (see Methods and Fig. 2e). On one hand, we found that the number of contacts was positively associated with individual-based education level in seniors (rate ratio RR = 1.16, 95% CI (1.08-1.25)) for high education level with respect to middle-low), while adults and young adults showed a similar number of contacts regardless of education level (RR = 0.99 (0.96-1.03) and RR = 0.95 (0.87-1.04), respectively, Fig. 2f). In a sensitivity analysis distinguishing by survey wave, we found that the association between number of contacts and education level was always positive for seniors, while for adults and younger individuals the sign of the association varied (Fig. S7). Moreover, household income was also found to be positively associated with the number of contacts (Fig. 2e), and the positive association was also found for all age groups after inclusion of an interaction term (Fig. S8). On the other hand, for some age groups we found a negative association between the number of contacts and SEP level. Adults with high SEP had fewer contacts with respect to low SEP (RR = 0.93 (0.90-0.97)). This effect was even stronger in young adults (RR = 0.84 (0.78-0.91)), while no significant effect was found in children and seniors (Fig. 2f). Similar results were found when restricting to contacts outside home (Fig. S6), and when stratifying by survey wave (Fig. S7). Overall, these results highlight heterogeneous contact activity across age and socio-economic groups, calling for the need to build a social contact matrix stratified by age, SEP, and education level.

### Expanded contact matrix

To build a fully stratified contact matrix, we developed a method to synthetically extend the available empirical data, as information on SEP and education level of contacts was missing. We first estimated an age-stratified contact matrix (Fig. 3a), adjusted for reciprocity, using a standard approach (see Methods). Then, we estimated an intermediate contact matrix stratified by SEP and education level on the participant side, using the information on education level and SEP provided in the survey (Fig. 3b). We applied a reciprocity correction to ensure consistency with the age-stratified contact matrix (see Supplementary Information). The cumulative number of contacts for each group relative to the total number of contacts is shown in Fig. 3c. Our method allows us to extend the intermediate contact matrix by distributing the age-specific number of contacts across the four missing socio-economic groups, while preserving conditions on aggregation and reciprocity of the expanded contact matrix (Methods). We used our method to generate 10,000 expanded contact matrices, which differ according to the choice of parameter values that define the level of assortativity across the socio-economic groups (see Methods and Supplementary Information for the definition and the interpretation of these parameters). One realization of a synthetic matrix generated through our method is displayed in Fig. 3d. We then characterised the properties of the generated contact matrices in terms of *R*_0_ and assortativity index, with respect to a matrix neglecting stratification by SEP and education level. For each expanded contact matrix, we computed the ratio of the dominant eigenvalue with respect to the age-only stratified contact matrix (equivalent to an expanded contact matrix assuming homogeneous mixing in the SEP and education level dimensions). We found that the ratio is always positive, meaning that including additional dimensions would increase the reproductive number *R*_0_ (as expected, see Methods and Ref.^34^). However, the estimated relative variation is relatively small, ranging between 0.5% and 1% (Fig. 3e). By aggregating over one of the two social dimensions, we found that, with respect to age-stratified mixing, the social dimension contributing the most to the relative change in the dominant eigenvalue (and therefore in *R*_0_ , assuming all groups are epidemiologically equivalent) is the education level (Fig. S14). With respect to a model with full homogeneous mixing, the relative variation in *R*_0_ obtained after introducing the age dimension is much higher than the variation obtained by integrating SEP or education level or the two combined (Fig. S15), suggesting that age is still the main driver of contact heterogeneity. For each expanded contact matrix, we computed two aggregated matrices, stratified only by SEP level (Fig. 3g) or by education level (Fig. 3h) and extracted an assortativity index, a summary measure of assortativity for each social dimension (Fig. 3f). On one hand, we found that, in the education level dimension, contacts can show both lower and higher assortativity with respect to homogeneous mixing (19% and 81% of cases, respectively). On the other hand, in the SEP dimension, contacts are almost always more assortative than homogeneous mixing (96% of cases), meaning that people with a given SEP level tend to have more contacts with people with the same SEP level. In both cases (SEP and education level), we found that, on average, the higher the assortativity, the higher the relative change in the dominant eigenvalue (Fig. S13). Overall, our method allows us to restrict the two-dimensional space of assortativity to values compatible with the observed empirical data.

**Figure 3.**
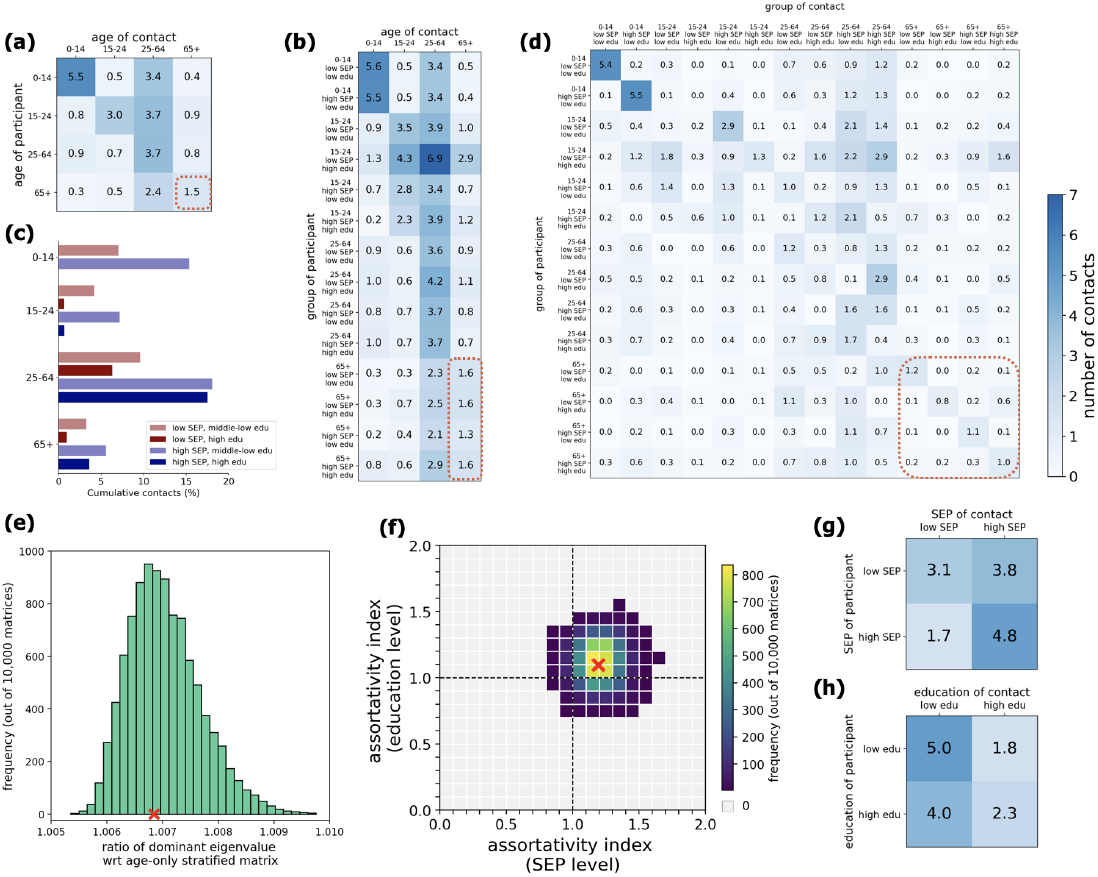
Expanded contact matrices. **(a)** Age-stratified contact matrix 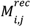, adjusted for reciprocity, informed from the available data. **(b)** Intermediate contact matrix 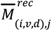 stratified by SEP and education level on the participants’ side, adjusted for reciprocity, informed from the available data. **(c)** Cumulative number of contacts (in %) engaged by a group, out of the total number of contacts, computed as 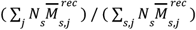 for each group *s* = (*i, v, d*) with age *i*, SEP level *v*, and education level *d*. **(d)** One example of a fully expanded synthetic contact matrix 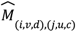, compatible with the age-stratified matrix *M*^*rec*^ and with the intermediate matrix 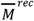 under aggregation. The matrix 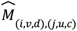 was obtained with a given combination of the free assortativity parameters (parameter values are reported in the Supplementary Information, Table S1). The dashed orange rectangles across panels (a), (b) and (d) indicate an example of matrix elements that are related through aggregation and stratification (in this case, mixing within the group of seniors 65+ year olds). **(e)** Distribution of the ratio of the dominant eigenvalue of the expanded contact matrices with respect to a matrix with homogeneous mixing in the SEP and education level dimensions. **(f)** Heatmap of the frequency of values of the assortativity index in the two dimensions (education level on y axis and SEP level on x axis). The assortativity index is defined so that a value equal to 1 indicates homogeneous mixing (i.e., mixing proportional to group sizes), and values larger and lower than 1 indicate more assortative and less assortative mixing, respectively, with respect to homogeneous mixing. Grey cells indicate ranges of assortativity not observed in our set of expanded contact matrices. In both panels (e) and (f), we show the results for 10,000 synthetic matrices. The values of assortativity index and dominant eigenvalue ratio for the example of expanded contact matrix shown in panel (c) are shown with a red cross in panel (e) and (f). **(g)** Matrix shown in (d) stratified by education level only. **(h)** Matrix shown in (d) stratified by SEP level only.

### Impact on epidemic spreading

We integrated our expanded contact matrices into an SIR epidemic model, to investigate how heterogeneous contact patterns along the education level and SEP dimensions can impact disease spreading. We considered two scenarios, namely (i) a scenario where all groups are epidemiologically equivalent, and (ii) a scenario where children (<15 year olds) have a reduced susceptibility with respect to other age groups (50% reduction), to mirror a COVID-19-like scenario^57^. First, to identify which groups are mostly affected by the disease, we computed the relative attack rate, i.e., the cumulative number of infections within each group. Second, to identify which groups contribute the most to transmission, we computed the relative contribution to *R*_0_ of each group, using the framework introduced in Ref.^53^. This index takes into account the total contact activity of each group, the mixing with other groups, and the sizes of the groups, together with their epidemiological characteristics (see Methods for additional details).

We found that, despite the small relative variation in *R*_0_ with respect to homogeneous mixing (Fig. 3e), accounting for additional social dimensions in the contact matrix allows to uncover heterogeneity in disease burden and contribution to transmission across groups (Fig. 4). Among the adult population (25-64 year olds), the most affected group consisted of individuals with high education and low SEP, who faced more infections than the average adults (Fig. 4a,c). This discrepancy is even stronger among young adults (15-24 year olds). Instead, among seniors (65+ year olds), the most affected group corresponded to individuals with a high education level living in high SEP areas. We found that the highest contribution to *R*_0_ was provided by individuals with high SEP, in particular children and adults in the scenario of homogeneous susceptibility (Fig. 4b), and adults and young adults in the scenario with reduced susceptibility for children (Fig. 4d). Therefore, even though *R*_0_ does not largely differ with respect to homogeneous mixing, the relative contribution and the role in transmission of each socio-economic group is highly heterogeneous.

**Figure 4.**
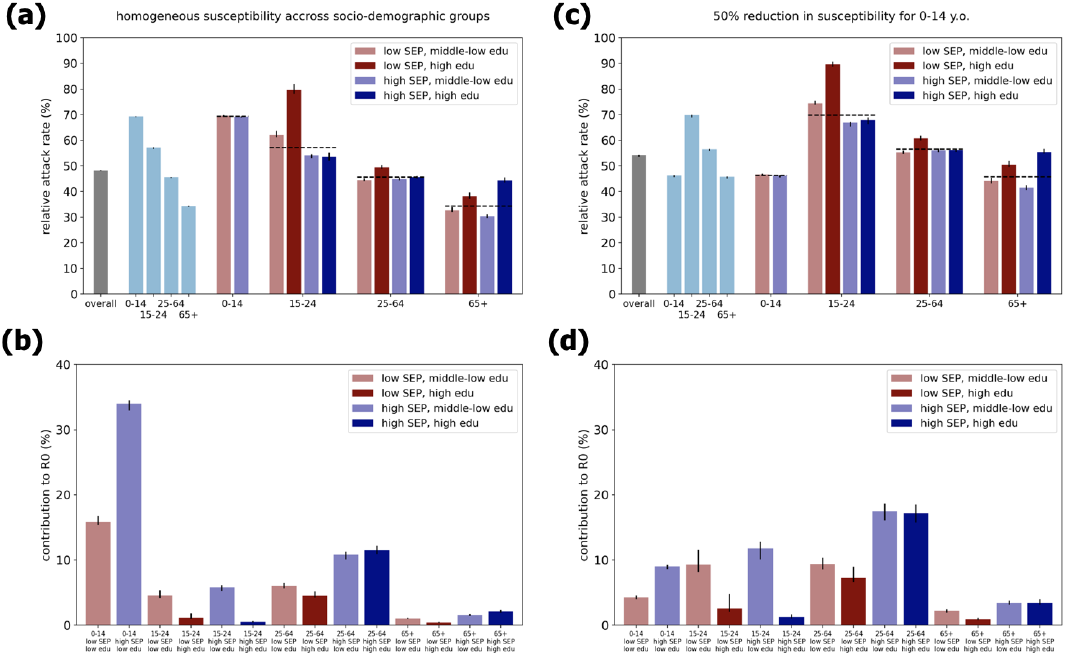
Epidemic spread in absence of control strategies. We considered a structured SIR epidemic model, stratified by age and socio-economic levels with heterogeneous mixing, and two epidemic scenarios: homogeneous susceptibility and infectiousness across age and socio-economic groups (panels a-b), or a reduction in susceptibility of 50% for children (panels c-d). In both scenarios, we assumed *R*_0_ = 1. 5, and an average infectious period of 3 days. **(a)** Relative attack rate for each group, i.e., fraction of the cumulative number of infected individuals over the size of the group. Color indicates the low SEP (red) and high SEP group (blue); color shade indicates groups with middle-low education level (lighter shade) and high education level respectively (darker shade). The attack rate overall (grey) and by age groups (light blue) are shown for comparison. Attack rates by age groups are also shown with vertical dashed lines for the corresponding socio-economic groups. **(b)** Proportional contribution to *R*_0_ of each group, computed using the framework of perturbation analysis of the next-generation matrix introduced in Ref.^53^. **(c-d)** As in (a-b), but assuming a lower susceptibility for the youngest age group of children (0-14 y.o) with a 50% reduction with respect to the other groups. In all panels, the bars represent median values; the whiskers represent the 2.5 and 97.5 percentiles computed across 1,000 realizations of the expanded synthetic contact matrices.

### Effectiveness of targeted control strategies

Next, we evaluated the effectiveness of control strategies. Here, we focused on targeted control strategies, i.e., strategies aiming at one group, while the rest of the population is unaffected. In a simplified scenario, we assumed that, at the start of the epidemic, a fraction of one targeted group *g* is permanently immune (e.g., through vaccination) or isolated due to a reduction in contacts. We determined whether there is a critical control effort (i.e., a critical fraction of the group computed as 1 − 1/*T*_*g*_ from the type reproduction number *T*_*g*_ , see Methods) which would prevent the spread of the epidemic in the full population. If so, we then computed the corresponding overall control effort defined as (1 − 1/*T*_*g*_ )*N_*g*_ /N where N_*g*_ is the population size of the group and N is the total population. The overall control effort thus represents the fraction of individuals targeted (belonging to the same group *g*) over the total population, and can be compared across strategies targeted at different groups. For each targeted strategy, we tested different expanded contact matrices with heterogeneous mixing, to investigate the interplay between assortativity and control effort. Given *R*_0_ = 1. 5, the estimated immunity threshold would be 33% under the assumption of a homogenous population (with random mixing).

We considered the epidemic scenario with reduced susceptibility for children, to model a disease similar to COVID-19^57^. Results for the homogeneous epidemic scenario are included in the Supplementary Information (Fig. S16). We tested different ways of partitioning the population. First, we grouped the population by SEP level (31% and 69% for low SEP and high SEP, respectively), or in another scenario by education level (69% and 31% for middle-low and high education, respectively). Sizes of the groups are reported in Fig. 5j. For the former partitioning, we found that targeting individuals with high SEP (the largest group) is successful in 31% of the cases (Fig. 5i), depending on the expanded contact matrix, and the overall control effort required would range between 34% and 69% (Fig. 5a). The lower the assortativity index in the SEP dimension, the lower the control effort required (Fig. 5c). When considering the individuals with low SEP as a target, we found very few contact matrices (1.2%) which would allow epidemic control. They correspond to very low assortativity levels in the SEP dimension (Fig. 5b), but if effective they would require a much lower control effort (27% to 31%, Fig. 5a). Similar considerations hold when targeting individuals with high or middle-low education (Fig. 5d), noting that in this case the largest group is represented by individuals with middle-low education (which include all children aged 0-14). The control strategy would be more effective (i.e., it would require a lower control effort) when contact patterns are similar to or less assortative than homogeneous mixing in the education level dimension (Fig. 5e,f), analogously to the results found for the SEP dimension. We then considered a partitioning in four groups as possible targets, i.e., individuals with low SEP and middle-low education, low SEP and high education, high SEP and middle-low education, high SEP high education (Fig. 5g,h). The type-reproduction number indicated that in this case only strategies targeted at the group with high SEP and low education (the largest group, accounting for 45% of the population, Fig. 5j) can be effective in controlling the epidemic. A strategy targeted to this group would be effective for 29% of the matrices considered (Fig. 5i). The overall control effort required ranges between 33% and 45% (Fig. 5g). From Fig. 5h, we found that a lower control effort would be needed when contacts are less assortative both in the SEP and education level, combining the effects observed with the previous independent strategies considering only education level or only SEP stratification. Overall, we found that the less assortative contacts are in the additional socio-economic dimensions (SEP and education level), the higher the chances that a targeted control strategy would be effective in preventing epidemic spread, and the lower the control effort needed.

**Figure 5.**
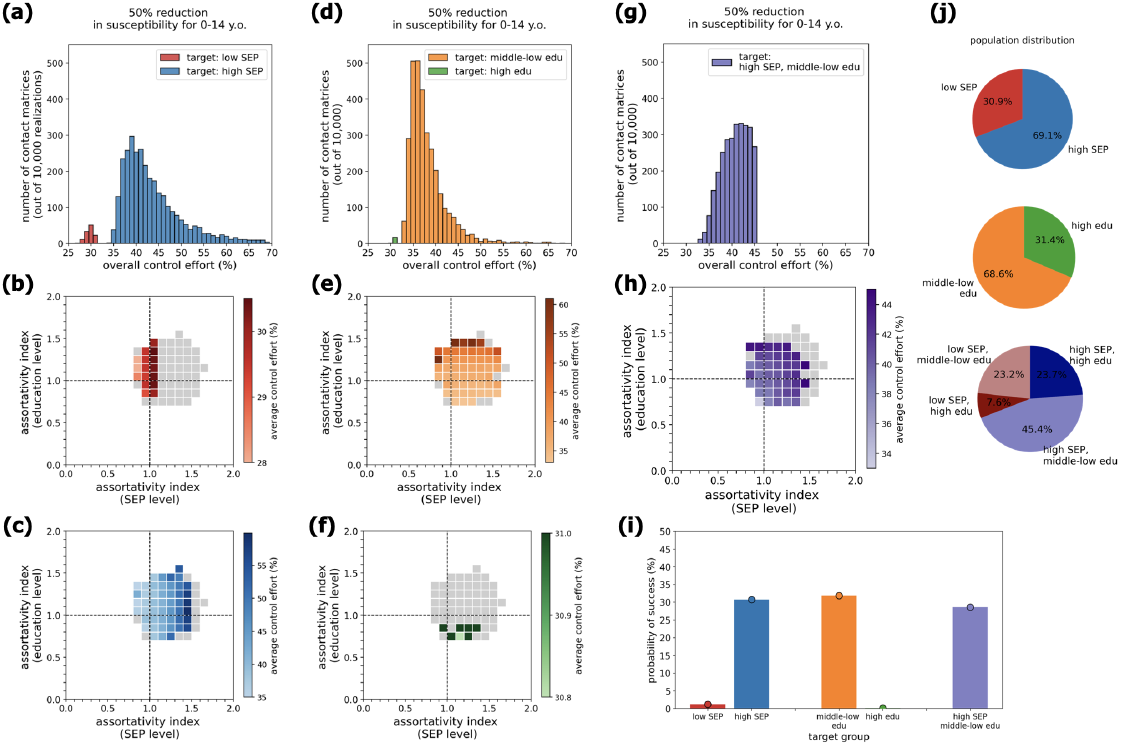
Effective targeted control strategies. Results for the epidemic scenario with reduced susceptibility for children. **(a)** Distribution of the overall control effort required by strategies targeted at individuals with low SEP (red) or with high SEP (blue). **(b)** Assortativity levels in the SEP and education dimensions for the subset of matrices which allow effective control (colored cells) and those matrices for which the strategy would not be effective (grey cells). The color gradient indicates the average control effort required for a given range of assortativity. The strategy considered here is targeted at the low SEP group. **(c)** As in panel (b), but considering a strategy targeted at the group with high SEP. **(c)** Distribution of the overall control effort required by strategies targeted at individuals with middle-low (orange) or with high education level (green). **(e)** As in panel (b), but considering a strategy targeted at the group with middle-low education. **(f)** As in panel (b), but considering a strategy targeted at the group with high education. **(g)** Distribution of the overall control effort required by strategies targeted at individuals with high SEP and middle-low education level. **(h)** As in panel (b), but considering a strategy targeted at the group with high SEP and middle-low education level. **(i)** Probability of success of the targeted strategy, defined as the fraction of contact matrices for which there exists a critical control effort allowing epidemic control. **(j)** Pie charts displaying the distribution of the population in three partitions, i.e., low SEP/high SEP (top), middle-low education/high education (center), and the combination of the two dimensions.

## Discussion

Social disparities observed in health-related outcomes highlight the need to integrate socio-economic heterogeneity in traditional age-stratified epidemic models^29–33^. In this study, we used survey data to characterize the number of contacts and mixing patterns across age and socio-economic groups, including both individual-based and neighbourhood-based socio-economic factors. We found that the number of contacts varied with education level and SEP level, depending on the age group. Despite missing socio-economic information on peers, we used information on survey participants to reconstruct contact matrices fully stratified by age, SEP, and education level, by varying the distribution of contacts across different socio-economic groups, while ensuring conditions on aggregation and reciprocity of the mixing matrix. This method allowed us to restrict the boundaries of contact assortativity along social dimensions, identifying ranges compatible with the observed contact data. By integrating the extended contact matrices into a transmission model, we identified the socio-economic groups that contribute the most to transmission and that experience the heaviest disease burden. We showed that contact assortativity determines the effectiveness of targeted control strategies, as higher levels of assortativity require larger control efforts to prevent an epidemic outbreak.

Regarding education level, and considering all contacts irrespective of the location, we found that seniors with higher education level reported a higher number of contacts with respect to seniors with middle-low education level. In a sensitivity analysis, this trend was detected both in adults and seniors when restricting to contacts engaged in locations outside the household. Also household income was positively associated with the number of contacts. We considered contact surveys collected in Switzerland in January, June, and December 2021. Our results are in line with studies from other countries showing that individuals with higher education level had a higher number of non-household contacts during the COVID-19 pandemic^9,10,58^, except for periods with more stringent measures where the opposite trend was observed^10,11^. The latter suggests that these socio-economic groups were better able to adapt to the epidemiological situation, by reducing and resuming their contacts according to non-pharmaceutical interventions^9^.

Besides individual-based socio-economic factors that are commonly taken into consideration when analysing social contacts, our work also considers a neighbourhood-based socio-economic factor (socio-economic position, SEP). We found that adults and young adults living in areas with a higher SEP reported fewer contacts than those living in low-SEP areas. This trend was consistent across survey waves and also robust for non-household contacts. In contrast with the positive association found for education level and income, these results suggest that neighbourhood-based socio-economic factors encode different information on contact behaviour with respect to individual-based factors. They also suggest that the relationship between socio-economic factors and number of contacts may depend on the age group, with individual-based factors (education level) being more relevant for seniors, and area-based factors (SEP) being more relevant for adults and young adults. This could be due to differences in lifestyle and habits. Area-based socioeconomic factors are often linked to mobility patterns^19,59^. Adults living in low-SEP areas may need to commute for reasons related to work, potentially engaging more contacts. In seniors, the number of contacts seems to be the same regardless of the SEP area, however seniors with higher education engage more contacts, potentially signaling the presence of larger social circles.

To integrate socio-economic factors into epidemic modeling, some studies used contact data to inform matrices stratified only on the participant side^9,60^, due to lack of socio-economic information about the contacts. The latter are often not collected through standard contact surveys, and would require detailed contact diaries where participants report additional information about contacts. The absence of these data prevents from uniquely identifying contact assortativity along socio-economic dimensions, and estimating an expanded matrix. Here, we presented a method to partially overcome this issue. Despite relying only on socio-economic information of the participants, we built a set of expanded contact matrices with varying assortativity levels, by exploiting matrix properties such as reciprocity and aggregation. Notably, we found that the assortativity levels compatible with the data do not span the full parameter space, and are actually rather constrained. This means that socio-economic stratification of participants, combined with information on the age for both participants and contacts, can provide some indication on the full mixing patterns. We found that the large majority of the expanded contact matrices exhibited a higher assortativity level than homogeneous mixing, for both individuals-based and neighbourhood-based socio-economic dimensions (education level and SEP). We found that assortativity was more pronounced for SEP than for education level. This could be due to household contacts sharing the same SEP, thus enhancing assortativity in this dimension. Overall, our results are in line with a study on contact data collected in Hungary during the COVID-19 pandemic, which showed preliminary evidence that contacts are assortative along the SES dimension (self-perceived wealth)^34^. Assortativity within socio-economic groups has also been observed in mobility patterns^61^.

Accounting for the number of contacts, heterogeneous mixing patterns and population sizes, we found that individuals contributing the most to transmission were children and adults living in high SEP areas. Integrating the matrices into a transmission model revealed heterogeneous disease burden across socio-economic groups, with higher attack rates in adults with high education living in low SEP areas and seniors with high education living in high SEP areas. Notably, these differences were found even under the assumption of equal conditions in susceptibility, infectiousness and disease severity across socio-economic groups. Other health-related heterogeneities, for example differences in vaccine uptake across socio-economic groups, could further exacerbate the differences in epidemic outcomes^35,62,63^.

The level of contact assortativity is an important factor in determining epidemic dynamics^64^. Using our reconstruction method, we were able to identify ranges of assortativity in education level and SEP dimensions, compatible with the aggregated contact data collected through the survey. We tested the impact of assortativity on the effectiveness of targeted control measures. We found that the lower the assortativity, the higher the chances of a targeted control strategy to be effective, and the lower the control effort needed to contain the epidemic (i.e., a smaller proportion of the total population needs to be targeted with immunization or contact reduction measures). Our results are in line with insights from epidemic spreading on networks^65–67^. Assortative networks are still prone to disease spreading despite immunization of part of the network, because they remain highly connected even after removal of some links. Instead, when contacts are less assortative, and closer to homogeneous mixing, effective epidemic control in a subgroup can be beneficial for the whole population, because individuals outside of the target group are less able to sustain the epidemic on their own.

Several modeling studies have accounted for socio-economic factors indirectly. For example, a COVID-19 modeling study in England used different age-stratified contact matrices for each area based on a spatial index of multiple deprivation, however matrices were only adapted for population structure, and did not account for different contact activity depending on area-based socio-economic level^60^. Our results suggest that integrating education level and SEP directly into the contact matrix can capture existing heterogeneities in contacts. In this perspective, modeling studies could also benefit from collection of epidemiological data (e.g., incidence and hospitalization) and other behavioral data (e.g., risk perception and mask wearing) stratified by socio-economic status and not only by age, to be integrated into transmission models together with contact data. Moreover, seroprevalence data could be used to fit the assortativity level of the contact matrix^68^.

We acknowledge some limitations of our study. First, participants in the survey did not report the exact neighbourhood of residence, but only the municipality. As an approximation, we assigned to each participant the average SEP index of the municipality of residence. Second, we pulled together data from three distinct survey periods, to reach an adequate sample size while stratifying into 16 groups (age, SEP and education level). We did not perform a longitudinal analysis looking at changes in contact behaviour over time, due to small sample sizes. Third, we used pandemic contact data and pooled participants’ responses over several survey waves, therefore our results may not be generalizable to pre- and post-pandemic behaviour. For Switzerland, we did not have pre-pandemic survey data to be used for comparison. A synthetic pre-pandemic age-stratified contact matrix for Switzerland was available^39,69^, but it was not stratified by socio-economic factors. Fourth, we considered illustrative control strategies, where only a subgroup of the population was targeted, while the rest of the population was not affected by public health measures. More complex interventions, while potentially more realistic, would have hindered the interpretation of the interplay between assortativity and control effort. Finally, we generated the expanded contact matrices by distributing the number of contacts across socio-economic groups of the peers, based on an intermediate contact matrix. The latter was estimated from the available data stratifying by age and socio-economic factors on the participants’ side. Therefore, any bias in these data could be reflected in the expanded contact matrices. More individual-level data would reduce potential biases and allow to generalize findings to the entire population.

In this study, we analysed contact behaviour in different socio-economic groups so far overlooked, and we developed a method to reconstruct a fully stratified contact matrix despite lacking socio-economic information about peers, providing insights on the interplay between contact assortativity and epidemic control. Future characterisation of post-pandemic contact behaviour accounting for additional socio-economic dimensions can help to inform more effective and equitable control measures to prevent infectious disease spread.

## Data Availability

Population and household statistics, and the dataset with the SEP index are available under request from external providers. All other population datasets used in this study are openly available. The social contact data, with a subset of variables for participants, are available on Zenodo; the full dataset can be obtained by the authors under request. The code used to run the analysis is available on Github at https://github.com/ISPMBern/comix_SEP.

## Data and code availability

The dataset with the SEP index can be requested here^70^. Population and household statistics (STATPOP^71^) 2021-2022 were provided by the Federal Statistical Office under user agreement for the scope of this project. All other population datasets used in this study are openly available from the Federal Statistical Office here^43,44^. The social contact data, with a subset of variables for participants, are available on Zenodo^40^; the full dataset can be obtained by the authors under request. The code used to run the analysis is available on Github at https://github.com/ISPMBern/comix_SEP.

## Author contributions

L.D.D. and C.L.A. conceived and designed the analysis. L.D.D. and M.L.R. cleaned and prepared the data. L.D.D. performed the analysis and wrote the manuscript. All authors critically revised the manuscript and approved its final version.

## Acknowledgments

This work has received funding from the ESCAPE project (101095619), funded by the European Union, and the Swiss State Secretariat for Education, Research and Innovation (SERI) (22.00482). The work was further supported by the Multidisciplinary Center for Infectious Diseases, University of Bern, Bern, Switzerland. The funders had no role in study design, data collection and analysis, decision to publish, or preparation of the manuscript.

## Ethics approval and consent to participate

The CoMix study protocols and questionnaires were approved by the local ethics committee of the Canton of Bern (project number 2020–02926). All methods were performed in accordance with regulations and informed consent of participants was obtained.

## Supplementary Information

## Data

### Survey participants and survey waves

The social contact data from Switzerland are presented in detail in another study^39^. To increase the sample size, we aggregated the adult participants from surveys categorized as A1, B1, and F1 (and for children, C1, D1 and E1), even though they refer to different periods, i.e., January - February 2021, June - July 2021 and December 2021 - January 2022, respectively. Survey periods are depicted in Fig. S1.

All participants in the survey reported the following information: age, gender, region (urban or rural), country of birth, education level, household income, employment status, household size, COVID-19 vaccination status, municipality of residence. We divided the participants into four age groups, i.e., 0-14, 15-24, 25-64, and 65+ year olds, to match available population data. We refer to these age groups as children, young adults, adults, and seniors, respectively. Age of children in the survey was reported in age-brackets. For simplicity, we assigned participants reported as 12-15 year olds to the age group of 0-14 year olds. For gender, we considered female, male, and other (the latter including answers such as ‘in another way’ or ‘prefer not to answer’). Country of birth was categorized as Switzerland, European Union, outside European Union, or unknown. We aggregated the education level into two categories: high education level, and middle-low education level. The former group corresponds to the tertiary level in the Swiss Education System^41^, which includes advanced education or a university degree (Bachelor, Master or PhD); the latter group includes individuals with upper-secondary education or without any post-compulsory education. All children were classified as middle-low education. Monthly household income was grouped in four categories: 0-5,000 CHF, 5,001-10,000 CHF, 10,000+ CHF, and ‘prefer not to answer’. Employment status was classified in two groups, i.e., employed or unemployed, with the latter including the following answers: ‘retired’, ‘student’, ‘homemaker’, ‘unemployed’, ‘other unemployed situation’. Household size was considered in three groups, as single units, couples or larger households with three or more members.

**Figure S1.**
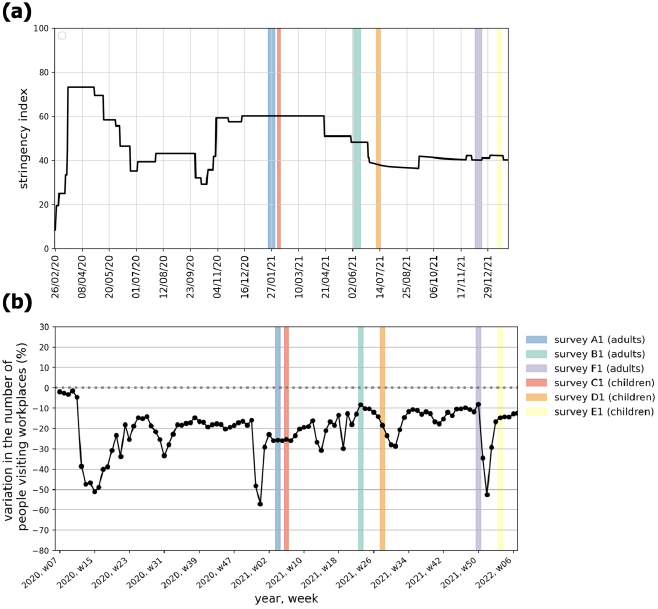
Pandemic context of the social contact surveys in Switzerland in terms of stringency index (Oxford COVID-19 Government Response Tracker^72^, panel (a)) and weekly mobility variation related to workplaces (Google mobility data^73^, panel (b)). In both panels, colored shaded areas refer to the period of data collection of the contact surveys.

### Weighted SEP index

The SEP index is defined for each residential building in Switzerland. Participants in the contact survey did not declare their address but only their municipality of residence. Therefore, to assign a SEP index to each participants, we computed a weighted average of the SEP index *wS*_*A*_ by municipality *A* as

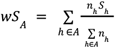

where *h* is any residential building in municipality *A, n*_*h*_ is the number of people living in residential building *h*, and *S*_*h*_ is the corresponding SEP index. In other words, we mapped the distribution of SEP values at residential building level to a distribution at individual level, and then computed the average SEP per individual living in the municipality. To inform *n*_*h*_ , we used the population and household statistic (STATPOP) 2021-2022 dataset^71^. We then used the median value of the weighted average SEP by municipality to classify participants in a given municipality as low or high SEP. Results are displayed in Fig. S2.

**Figure S2.**
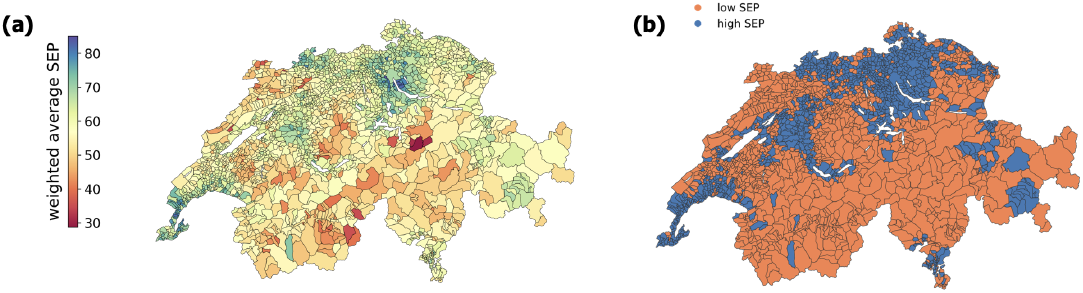
Weighted SEP index by municipality. **(a)** Map of municipalities in Switzerland color-coded based on the value of the weighted SEP. **(b)** Map of municipalities in Switzerland classified as low SEP (orange) or high SEP (blue) using the median weighted SEP as a threshold.

### Population size by age group, SEP level and education level

Participants in the CoMix survey could be directly classified in terms of age group, SEP level (based on the municipality of residence) and education level based on the information available (see section above). For the general Swiss population, we extracted an estimate of population sizes using the following procedure.

We used data by education level (according to the 3-group classification used by the Swiss Education System, i.e., without post-compulsory education, upper-secondary level, tertiary level) available for each district (an administrative area coarser than municipalities) for the permanent adult population (above 25 y.o.). For each district, we computed the population profile by education level, and we assigned the weighted SEP index of the district (computed analogously to the weighted SEP index by municipality). For each district, we also computed the population size of the four age groups of interest (0-14, 15-24, 25-64, 65+ y.o.). To infer the missing interaction between age and education level in each district, we projected the distribution by education level and age observed at national level to the sizes of the groups by age and by education level for each district.

Quotas in the recruitment of survey participants for the CoMix were set on age and sex, allowing for a representative distribution in the sample along these variables, but not on education level and SEP. In Fig. S3 we checked for the representativeness of the sample including education level and SEP. We found good agreement between the sample and the general population.

**Figure S3.**
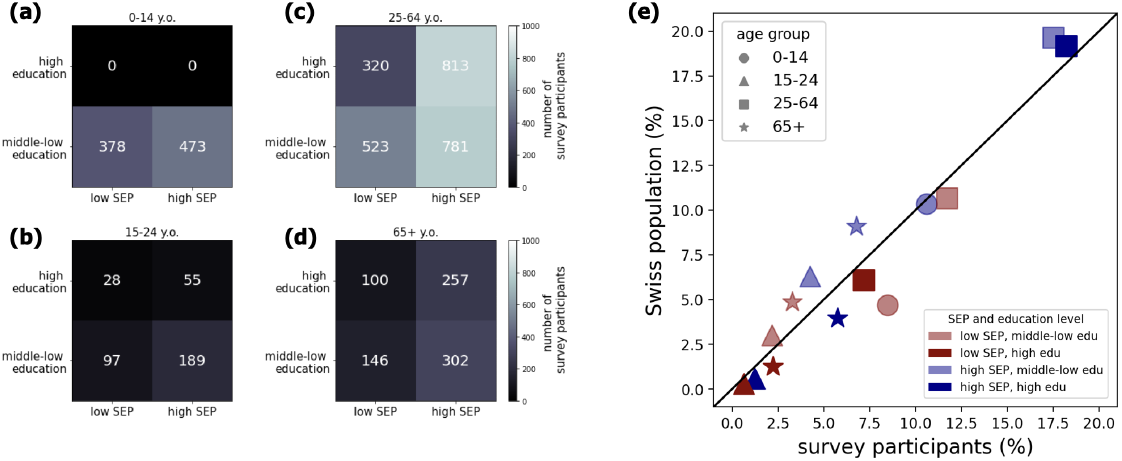
Representativeness of survey population. **(a)** Number of participants 0-14 y.o. divided by education level (y axis) and SEP level (x axis). **(b-d)** As in panel (a), showing the number of participants in age groups 15-24, 25-64 and 65+ y.o., respectively.**(e)** Population profile by age, SEP and SES, in Switzerland (y axis) and in the survey (x axis).

### Contact activity by survey wave

For completeness, we report in this section the results analogous to Fig. 2a-d in the main text, stratified by survey wave (Fig. S4).

**Figure S4.**
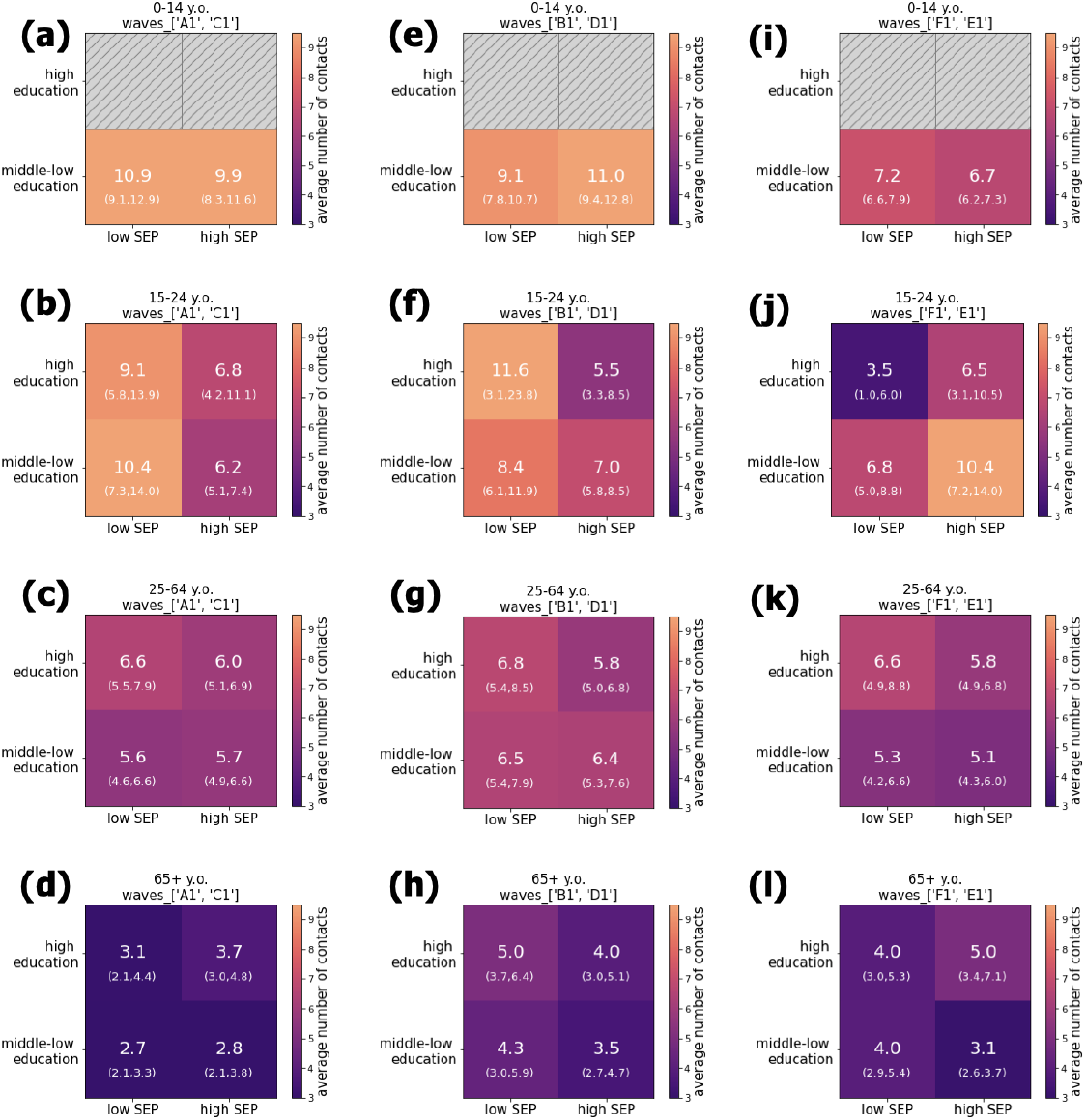
**(a-d)** Contact activity by individual-based (education level) and area-based (SEP) groups. The heatmap shows the crude mean number of contacts (along with 95% confidence interval obtained with 1000 bootstraps) engaged by a participant belonging to one of the four socio-economic groups, depending on the age group **(a)** 0-14 y.o. (children), **(b)** 15-24 y.o. (young adults) **(c)** 25-64 y.o. (adults) and **(d)** 65+ y.o. (seniors). Results refer to survey waves A1 and C1 (January - February 2021). **(e-h)** As in panels (a-d), results refer to survey waves B1 and D1 (June - July 2021). **(i-l)** As in panels (a-d), results refer to survey waves F1 and E1 (December 2021 - January 2021).

### Regression model

#### Sensitivity analyses

In this section, we present the results of some model variations in the regression analysis. In particular, we tested the inclusion of an additional explanatory variable (population density) and a different definition of the outcome variable, where we looked specifically at contacts at work and contacts outside the household rather than contacts overall. For contacts outside the household, we tested two definitions: contacts with a known location different from ‘home’ or ‘household’ (excluding contacts with missing location), or any contact outside home (including contacts with missing location). We also report results of the sensitivity analysis adding an interaction term with age for the household income variable, and distinguishing by survey wave.

For what concerns contacts at work, the number of contact was very low for all age groups except 25-64 y.o. (as expected). Therefore, the estimated rate ratios were either non significant or highly uncertain. For adults in 25-64 y.o., we found a negative effect of high SEP and high education level on the number of contacts at work, but the effect was very small (Fig. S5a).

For contacts outside home (in both definitions excluding or including unspecified locations, Fig. S5b,c), we found again a positive effect of high education level on the number of contacts in seniors (65+ y.o.) and a negative effect of SEP on the number of contacts in adults (25-64 y.o.), consistently to what was found in the main analysis on overall contacts.

We computed the population density (by km2) as the population divided by the area (km2). The area was retrieved by the polygons in the shapefile of Switzerland^74^, using an appropriate projection (Swiss Oblique Mercator, EPSG:2056). However, this measure does not account for large non-inhabited areas in Switzerland (due to e.g. mountains), especially in the south of the country. This is visible in the map shown in Ref.^37^. This means that a municipality which overlaps with a mountain area may have a low population density which is not representative of the effective population density in the inhabited areas. To correct for this bias, we computed the effective population density, as a weighted mean of the population by hectare. We used publicly available data on the population by hectare^75^.

We included the effective population density as a variable in the regression model, categorized as ‘low’, ‘baseline’ or ‘high’, using quartiles at 25% and 75% as threshold. We found that the estimates of the rate ratio did not change significantly. Results are shown in Fig. S6b.

Stratifying by survey wave, we found that the main results remained consistent across waves: seniors with higher education level reported more contacts with respect to middle-low education, and adults living in high SEP areas reported less contacts with respect to low SEP areas (Fig. S7).

In the main analysis, we found that household income was also positively associated with the number of contacts. This holds for all age groups after inclusion of an interaction term with age. The effect is smaller in adults and larger in young adults and seniors (Fig. S8b).

**Figure S5.**
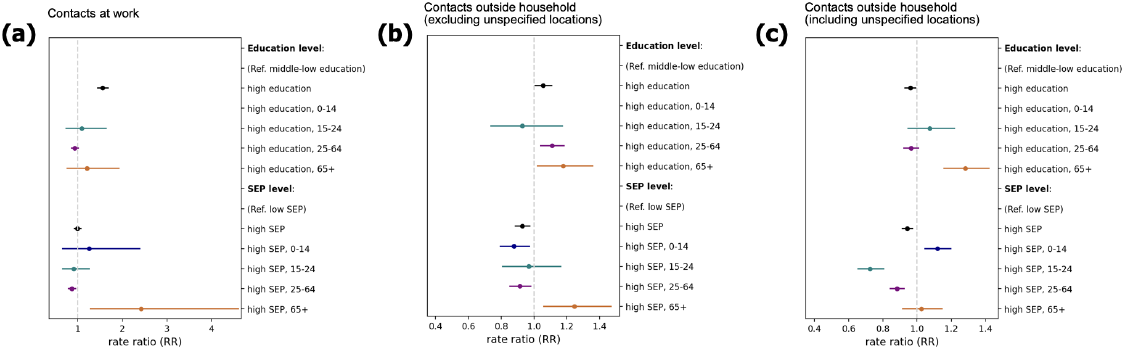
Changing the type of contacts in the outcome variable. **(a)** Same panel as the one shown in Fig. 2e in the main text, but using the number of contacts at work as the outcome variable. **(b)** As in panel (a), but using the number of contacts outside home (excluding unspecified locations) as outcome variable. **(c)** As in panel (a), but using the number of contacts outside home (including unspecified locations) as outcome variable. Black and colored indicate the rate ratio (RR) in the univariate and multivariate regression model with interaction term with age, respectively. Bars indicate 95% confidence intervals.

**Figure S6.**
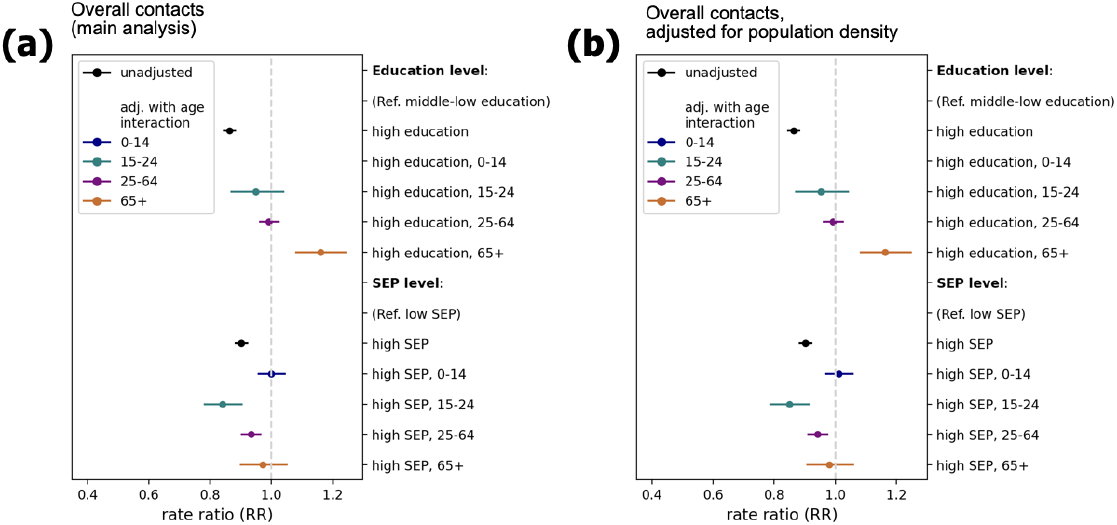
Adding population density as an explanatory variable. **(a)** Same panel as the one shown in Fig. 2e in the main text. Black and colored indicate the rate ratio (RR) in the univariate and multivariate regression model with interaction term with age, respectively. Bars indicate 95% confidence intervals. **(b)** As in panel (a), but adding the effective population density as a variable in the regression model.

**Figure S7.**
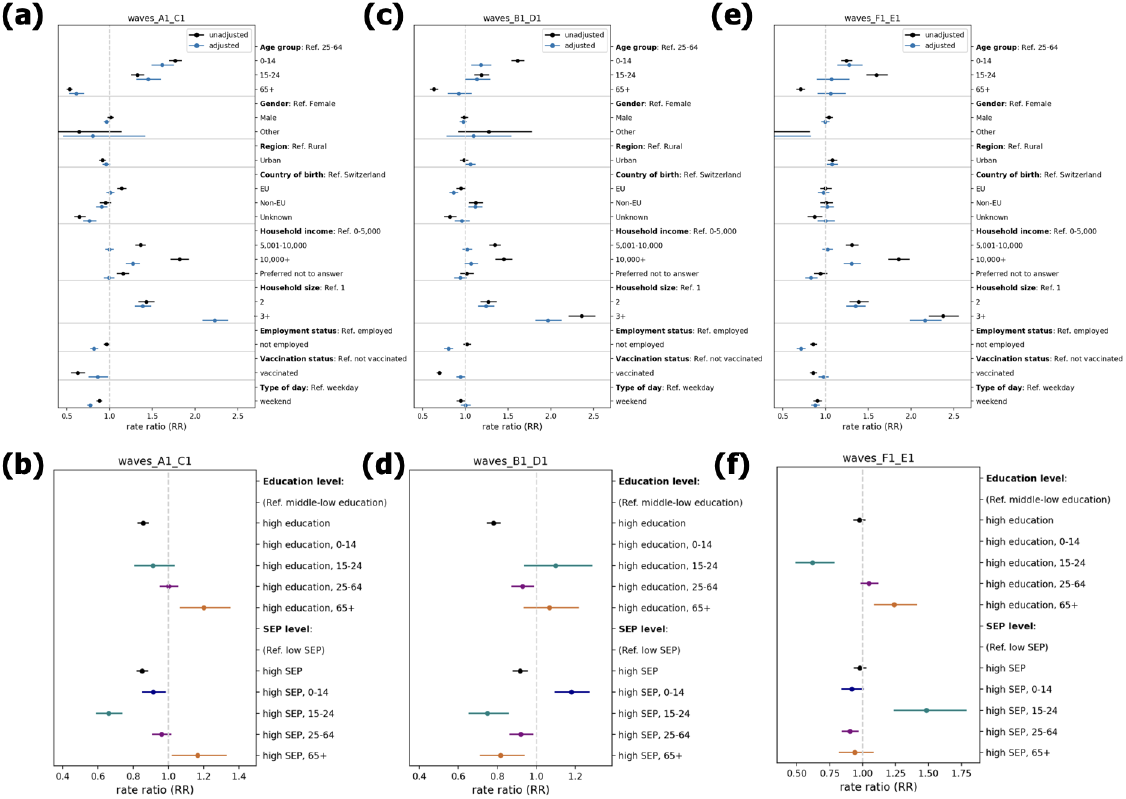
Stratifying by survey wave. Same panels as the ones shown in Fig. 2e,f in the main text, however considering only participants in the first survey period (January and February 2021 for adults and children respectively) in panels (a-b), in the second survey period (June and July 2021) in panels (c-d), and in the third survey period (December 2021 and January 2022) in panels (e-f). Black and colored indicate the rate ratio (RR) in the univariate and multivariate regression model with interaction term with age, respectively. Bars indicate 95% confidence intervals.

**Figure S8.**
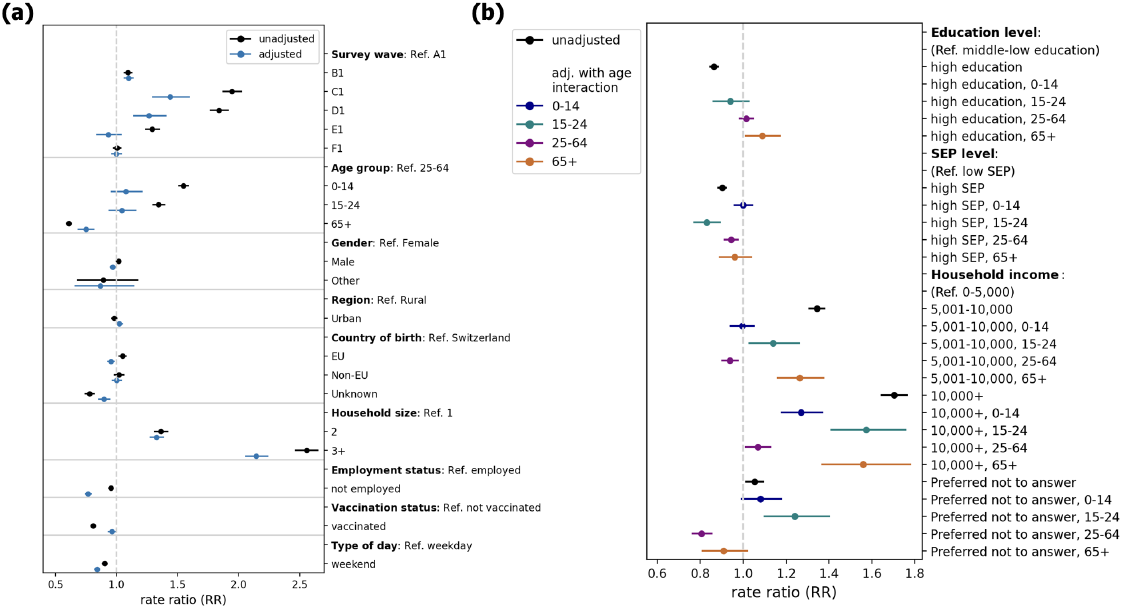
Adding an interaction term between age and income. Same panel as the ones shown in Fig. 2e,f in the main text, but including an interaction term not only for SEP and education level but also for household income. Black and colored indicate the rate ratio (RR) in the univariate and multivariate regression model with interaction term with age, respectively. Bars indicate 95% confidence intervals.

### Contact matrices

#### Reciprocity correction

In this section, we follow the notation introduced in the main text. We want to apply a reciprocity correction to the rectangular matrix 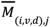,whose elements represent the number of contacts that a participant in age group *i*, SEP level *v* and education level *d* has with individuals in age group *j*. We applied a similar method as the one introduced in Ref.^9^. For each couple of age groups *i* and *j*, we computed 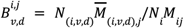 i.e the proportion of contacts engaged by individuals in socio-economic group (*v, d*) out of the total number of contacts between age group *i* and age group *j*.

We then applied these proportions to the elements of the age-stratified reciprocal matrix 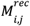 , to obtain 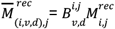, so that 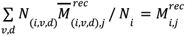. In other words, this correction ensures that, when aggregating the matrix 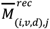 over the socio-economic groups (*v, d*), we can retrieve the reciprocal age-stratified matrix 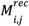

### Matrix expansion

We expanded the adjusted intermediate matrix 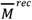 (Fig. S9a) to a fully stratified contact matrix 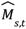 (Fig. S9b). To derive the elements of the matrix 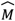, we shall solve a system of linear equations, with constraints due to the properties of the structure of the social contact matrix. In particular, the matrix 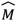 is required to fulfill conditions on (i) reciprocity, i.e., the total number of contacts 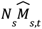 between individuals in group *s* and group *t* must be equal to 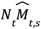; (ii) aggregation, i.e., the number of contacts summed over all socio-economic groups (excluding age) must be consistent with the elements of the intermediate contact matrix 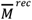 , so that 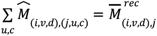 ; (iii) positivity, i.e., the number of contacts in each cell cannot be negative.

Practically, the global system can be solved through 10 independent linear systems corresponding to specific blocks of the contact matrix: 4 diagonal 4×4 blocks (one for each age group) with 16 variables, and 6 couples of off-diagonal 4×4 blocks, for a total of 32 variables, corresponding to couples of interacting age groups (*i, j*) with *i* ≠ *j*. In order to solve the system for each diagonal block (mixing within an age group), it is required to set 6 free parameters, while the remaining 10 are derived from the conditions on reciprocity and aggregation. We defined 6 parameters *q ∈* (0, 1), that can be interpreted as assortativity parameters along the education level dimension and the SEP dimension. To solve the system for the off-diagonal blocks (mixing across age groups), we found that it is required to set 9 free parameters, while the remaining 23 are derived from reciprocity and aggregation.

Similarly to above, we defined 9 parameters describing the assortativity and the general distribution of contacts along the education level and SEP dimension. In total, to derive the expanded, it is required to set the value of 55 free parameters. In Table S1, we report the values of the 55 parameters chosen for the matrix displayed in Fig. 3c (here also reported in Fig. S9b).

We explored the parameter space through random sampling in the range (0, 1) for each *q*, and selected combinations of parameter values that ensure the positivity of each block, and therefore of the expanded contact matrix. We exploited some analytical conditions on some parameters to restrict the parameter space and optimize the search. Additional details are contained in the following two sections.

**Figure S9.**
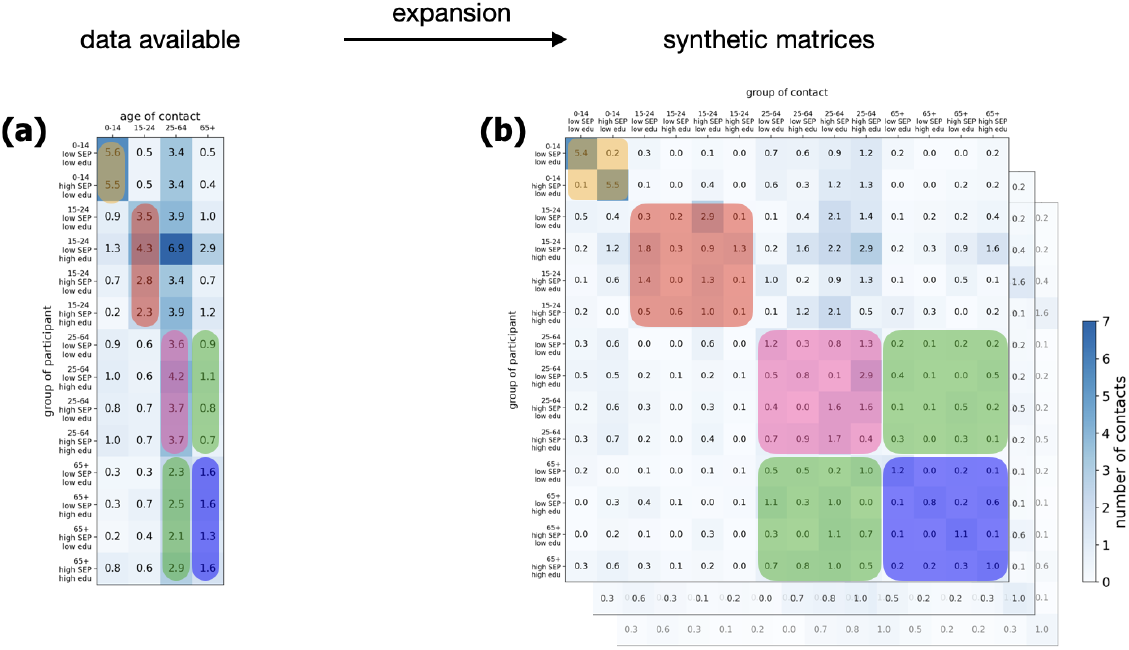
Scheme of the correspondence between the elements in the intermediate matrix (panel (a)) and the elements of the fully stratified matrix (panel (b), one example of the set of possible matrices). Some blocks are highlighted in color for illustrative purposes. The 4×4 diagonal block in pink represents the mixing within the age group of adults (25-64 y.o.), across the 4 socio-economic groups (low/high SEP, middle-low/high education), which must be compatible under aggregation (sum per row) with the elements in pink in the intermediate matrix. The off-diagonal block in green represents the mixing between adults (25-64 y.o.) and seniors (65+ y.o.), compatible under aggregation with the elements in green in the intermediate matrix. The matrix in panel (b) is built in order to satisfy reciprocity, i.e., *N*_*s*_*M*_*s*,*t*_ = *N*_*t*_*M*_*t*,*s*_ for any group *s, t* (3-tuple of age group *i*, SEP level *v* and education level *d*).

**Table S1.** List of parameter values used to generate the example of an expanded synthetic matrix shown in Fig. 3c (and Fig. S9b). For the definition of the parameters, see the sections below. The indexes *v*_1_ and *v*_2_ refer to the two SEP levels (low SEP and high SEP, respectively). The indexes *d*_1_ and *d*_2_ refer to the two education levels (middle-low and and high education level, respectively). For children, the parameters assortativity along the education level dimension are not defined because all children are classified as middle-low education.

| Mixing within an age group (diagonal blocks) |  |  |  |  |  |  |  |  |  |
| --- | --- | --- | --- | --- | --- | --- | --- | --- | --- |
| Age group $i$ | $q_{v_1, d_1}^i$ | $q_{v_1, d_2}^i$ | $q_{v_2, d_1}^i$ | $q_{v_2, d_2}^i$ | $q_{v_1}^i$ | $q_{d_1}^i$ | | | |
| Children | //////// | //////// | //////// | //////// | 0.97 | //////// |  |  |  |
| Young adults | 0.09 | 0.08 | 0.45 | 0.06 | 0.17 | 0.94 |  |  |  |
| Adults | 0.34 | 0.19 | 0.44 | 0.11 | 0.37 | 0.56 |  |  |  |
| Seniors | 0.79 | 0.50 | 0.80 | 0.62 | 0.74 | 0.90 |  |  |  |
| Mixing across two age groups (off-diagonal blocks) |  |  |  |  |  |  |  |  |  |
| Age groups $i, j$ | $q_{v_1, d_1}^{i, j}$ | $q_{v_1, d_2}^{i, j}$ | $q_{v_2, d_1}^{i, j}$ | $q_{v_2, d_2}^{i, j}$ | $q_{v_1}^{i, j}$ | $q_{d_1}^{i, j}$ | $r_1^{i, j}$ | $r_2^{i, j}$ | $r_3^{i, j}$ |
| Children & young adults | 0.65 | //////// | 0.71 | //////// | 0.67 | //////// | //////// | //////// | //////// |
| Children & adults | 0.20 | //////// | 0.35 | //////// | 0.37 | //////// | //////// | //////// | //////// |
| Children & seniors | 0.51 | //////// | 0.36 | //////// | 0.52 | //////// | //////// | //////// | //////// |
| Young adults & adults | 0.02 | 0.23 | 0.26 | 0.12 | 0.15 | 0.59 | 0.11 | 0.41 | 0.51 |
| Young adults & seniors | 0.11 | 0.12 | 0.69 | 0.16 | 0.28 | 0.65 | 0.22 | 0.33 | 0.52 |
| Adults & seniors | 0.29 | 0.06 | 0.63 | 0.14 | 0.46 | 0.64 | 0.22 | 0.04 | 0.63 |

#### Inferring matrix elements in the diagonal blocks

For each of the four diagonal blocks (one for each age group), composed of 16 unknowns, we can write down 10 conditions, namely 4 conditions for the aggregation and 6 conditions on the reciprocity. We then defined 6 conditions based on 6 free parameters that describe how contacts within the same age group are distributed across different SEP and education levels. More specifically, we defined one assortativity parameter for each of the 4 socio-economic groups (couple (*v, d*) of SEP and education level), one assortativity parameter in the SEP dimension only, and one assortativity parameter in the education level dimension only.

Given one age group *i*, we can define four parameters 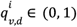 for any couple of SEP level *v ∈* {*v*_1_, *v*_2_} and education level *d ∈* {*d*_1_, *d*_2_} as the proportion of contacts engaged within the same group, i.e., 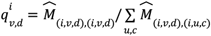 , where 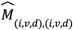 are the diagonal elements of the matrix 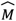.

We can then define the assortativity parameter along the SEP dimension and education level dimension separately by looking at the aggregated squared matrices 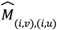 and 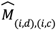 expanded only on the SEP level and age, or education level and age, respectively, and the corresponding rectangular matrices 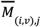 and 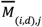 aggregated over the socio-economic group of the contactee. In the education level dimension, let us call *d*_1_ the group with middle-low education, and *d*_2_ the group with high education. We can define 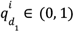 as

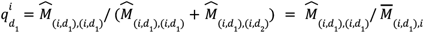

i.e., the share of contacts that one individual in education level *d*_1_ and age group *i* engages with the same group, regardless of the SEP level. Analogously, we can define 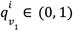 using *v*_1_ and *v*_2_ for the low SEP and high SEP groups.

For each age group *i*, we can then numerically solve the system of 16 equations for any combination of values for the six free parameters 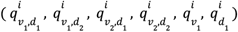 . In principle, any assortativity parameter *q* defined above could take any value between 0 and 1, by construction. However, not all values would be compatible with the condition of positivity, which we have not used so far.

For example, for 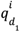 we can write down the following system

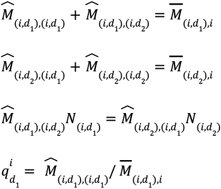

where 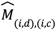 with *d, c ∈* {*d*_1_ , *d*_2_ } are the four unknown variables, while 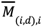 and 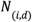 are known from the data. The first two conditions satisfy the aggregation property, while the third ensures reciprocity. By analytically solving this system, we can derive the following condition on 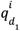:

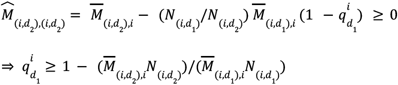

This condition is necessary (but not sufficient) in order for 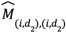 to be non-negative.

We can derive an analogous condition for 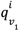 by looking at the 2×2 diagonal blocks of the matrix 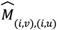 . Hence, when exploring the parameter space 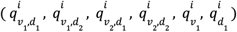 , we can restrict the values for 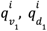 to the ones displayed in Table S2.

**Table S2.** Conditions on assortativity in the education level or SEP dimension necessary for positivity of the matrix elements in the four 4×4 diagonal blocks. The parameter 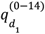 is equal to 1, because all children are classified as middle-low education, therefore all contacts are engaged within this group.

|  | SEP | Education level |
| --- | --- | --- |
| Children (0-14 y.o.) | $0 \leq q_{v_1}^{(0-14)} \leq 1$ | $q_{d_1}^{(0-14)} = 1$ |
| Young adults (15-24 y.o.) | $0 \leq q_{v_1}^{(15-24)} \leq 1$ | $0.90 \leq q_{d_1}^{(15-24)} \leq 1$ |
| Adults (25-64 y.o.) | $0 \leq q_{v_1}^{(25-64)} \leq 1$ | $0.13 \leq q_{d_1}^{(25-64)} \leq 1$ |
| Seniors (65+ y.o.) | $0 \leq q_{v_1}^{(65+)} \leq 1$ | $0.55 \leq q_{d_1}^{(65+)} \leq 1$ |

We can interpret the values in the table as follows. For example, the condition 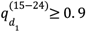 means that, out of the number of contacts that an individual with age (15-24 y.o.) and middle-low education level has on average with people in the (15-24 y.o.) group, at least 90% must be shared with people with the same middle-low education level.

We could in principle derive some conditions on the parameters 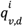 as well, however that would entail to analytically solve the system in 16 variables, and would require more cumbersome computations. Hence, we will check for values ranging between 0 and 1 through random sampling from a uniform distribution. In practice, we applied the following procedure. After a preliminary exploration, we identified some empirical constraints, i.e., some threshold values limiting the observed distribution. We used them in a second round of exploration, where we tested 2,000,000 parameter combinations by sampling each parameter from a uniform distribution over the corresponding interval, either (0-1) or a restricted interval based on the empirical or analytical constraints. We selected only those combinations leading to a positive contact matrix. The resulting selected distributions are illustrated in Fig. S10. For each parameter, we show the median value and the value expected from proportional mixing. We can see that in the majority of cases matrices display assortativity values higher than the ones expected from homogeneous mixing.

**Figure S10.**
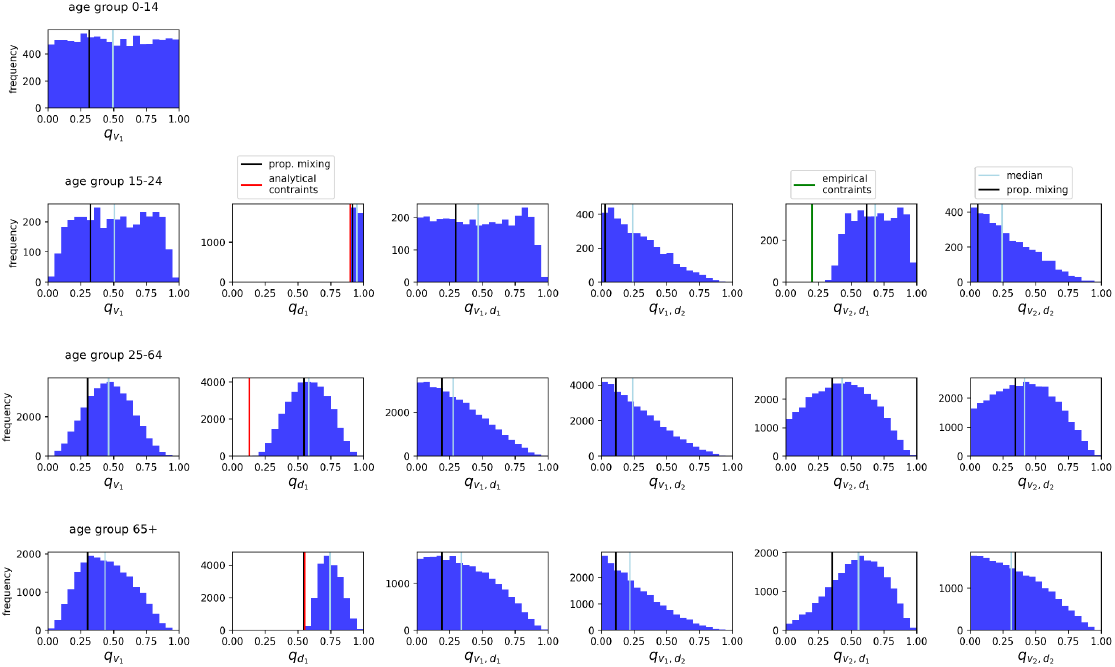
For each age group of young adults 15-24 y.o. (second row), adults 25-64 y.o. (third row) and seniors 65+ y.o (fourth row), we show the frequency of values of the 6 free parameters (columns) of assortativity which allow solving the system and derive a positive matrix. For children in 0-14 y.o., the system of equations is reduced because all children belong to the middle-low education group, therefore, only one parameter of assortativity 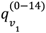 is needed (first row). In each panel we show the median of the observed distribution (vertical line in light blue), and the value expected from homogeneous mixing, i.e., mixing proportional to the size of the subgroups (vertical black line).

#### Inferring matrix elements in the off-diagonal blocks

For each of the six coupled off-diagonal blocks with 32 variables, corresponding to a couple of age groups *i* and *j* with *i* ≠ *j*, we can write 16 conditions on reciprocity, and 8 conditions on the aggregation. However, it turns out that one of them is linearly dependent on the others, so we can drop one condition on reciprocity and then set 9 remaining conditions in order to solve the system. Similarly to what was done before, we can define 9 free parameters which describe how contacts are distributed along the SEP and education level dimensions. We choose to fix: (i) 4 conditions on the assortativity on the (sub)diagonal elements; (ii) 2 conditions on the assortativity in the education level and SEP dimension separately, similarly to what was done above; (iii) 3 remaining parameters with no particular link with assortativity, defined on the share of contacts in one element outside of the sub-diagonal elements. More precisely, for a couple of age groups *i* and *j*:

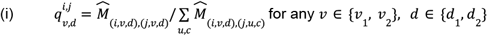

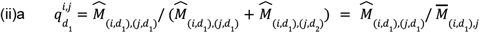

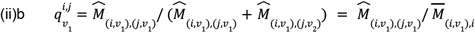

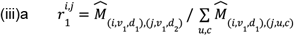

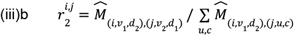

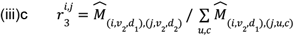

Similarly to what was done above, we can analytically solve the system for the blocks of the contact matrix expanded only on the education level and age, or only on the SEP level and age. We find that a necessary condition for the positivity is:

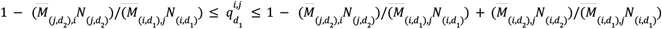

and an analogous condition can be written for 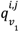 . Hence, when exploring the parameter space 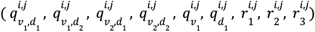 , we can restrict the values for 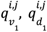 to the ones displayed in Table S3.

**Table S3.** Conditions on assortativity in the education level or SEP dimension necessary for positivity of the matrix elements in the three 4×4 diagonal blocks.

|  | SEP | Education level |
| --- | --- | --- |
| Young adults & adults | $0 \leq q_{v_1}^{(15-24),(25-64)} \leq 0.77$ | $0.49 \leq q_{d_1}^{(15-24),(25-64)} \leq 0.62$ |
| Young adults & seniors | $0 \leq q_{v_1}^{(15-24),(65+)} \leq 0.62$ | $0.53 \leq q_{d_1}^{(15-24),(65+)} \leq 0.75$ |
| Adults & seniors | $0 \leq q_{v_1}^{(25-64),(65+)} \leq 0.94$ | $0.43 \leq q_{d_1}^{(25-64),(65+)} \leq 1$ |
| Children & young adults | $0 \leq q_{v_1}^{(0-14),(15-24)} \leq 1$ | $0.93 \leq q_{d_1}^{(0-14),(15-24)} \leq 1$ |
| Children & adults | $0 \leq q_{v_1}^{(0-14),(25-64)} \leq 0.99$ | $0.51 \leq q_{d_1}^{(0-14),(25-64)} \leq 1$ |
| Children & seniors | $0 \leq q_{v_1}^{(0-14),(65+)} \leq 0.76$ | $0.44 \leq q_{d_1}^{(0-14),(65+)} \leq 1$ |

We can interpret the values in the table as follows. For example, the condition 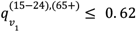 means that at maximum 62% of the number of contacts engaged by an individual with age (15-24 y.o.) and low SEP level with 65+ y.o. is shared with people with the same low SEP level.

Similarly to what was done for the diagonal blocks, we explored the 9-dimensional parameter space through random sampling. In practice, we applied the following procedure. After a preliminary exploration, we identified some empirical constraints, i.e., some threshold values limiting the observed distribution. We used them in a second round of exploration, where we tested 2,000,000 parameter combinations by sampling each parameter from a uniform distribution over the corresponding interval, either (0-1) or a restricted interval based on the empirical or analytical constraints.

We selected only those combinations leading to a positive contact matrix. The resulting selected distributions are illustrated in Fig. S11. For children in (0-14), the system is reduced to 8 equations because they all belong to the group with middle-low education, therefore the number of free parameters needed is 3 rather than 9. They are shown in Fig. S12.

**Figure S11.**
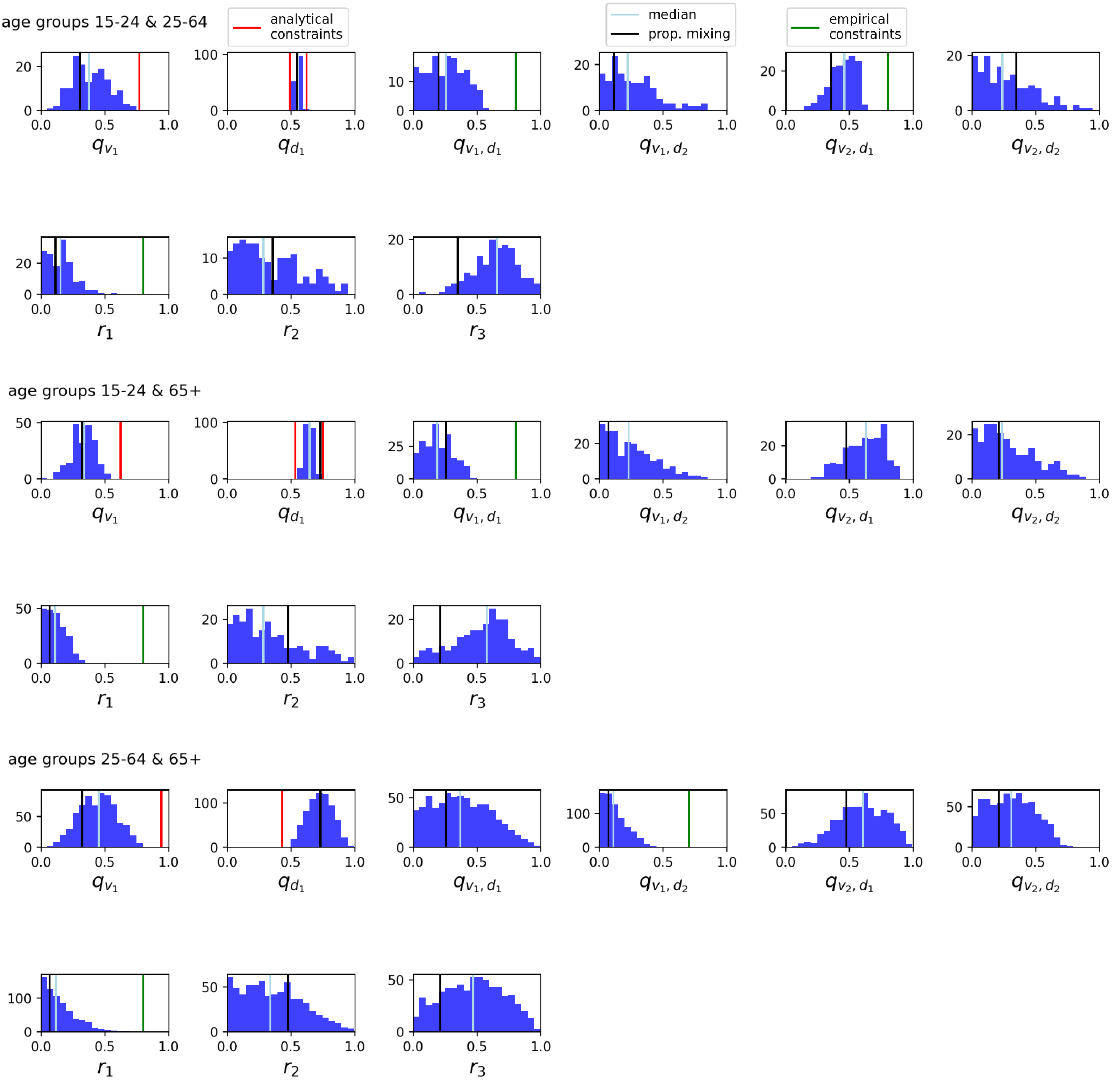
Frequency of values of the 9 free parameters in the selected combinations, for each couple of interacting age group (off-diagonal blocks), excluding children. In each panel we show the median of the observed distribution (vertical line in light blue), and the value expected from homogeneous mixing, i.e., mixing proportional to the size of the subgroups (vertical black line).

**Figure S12.**
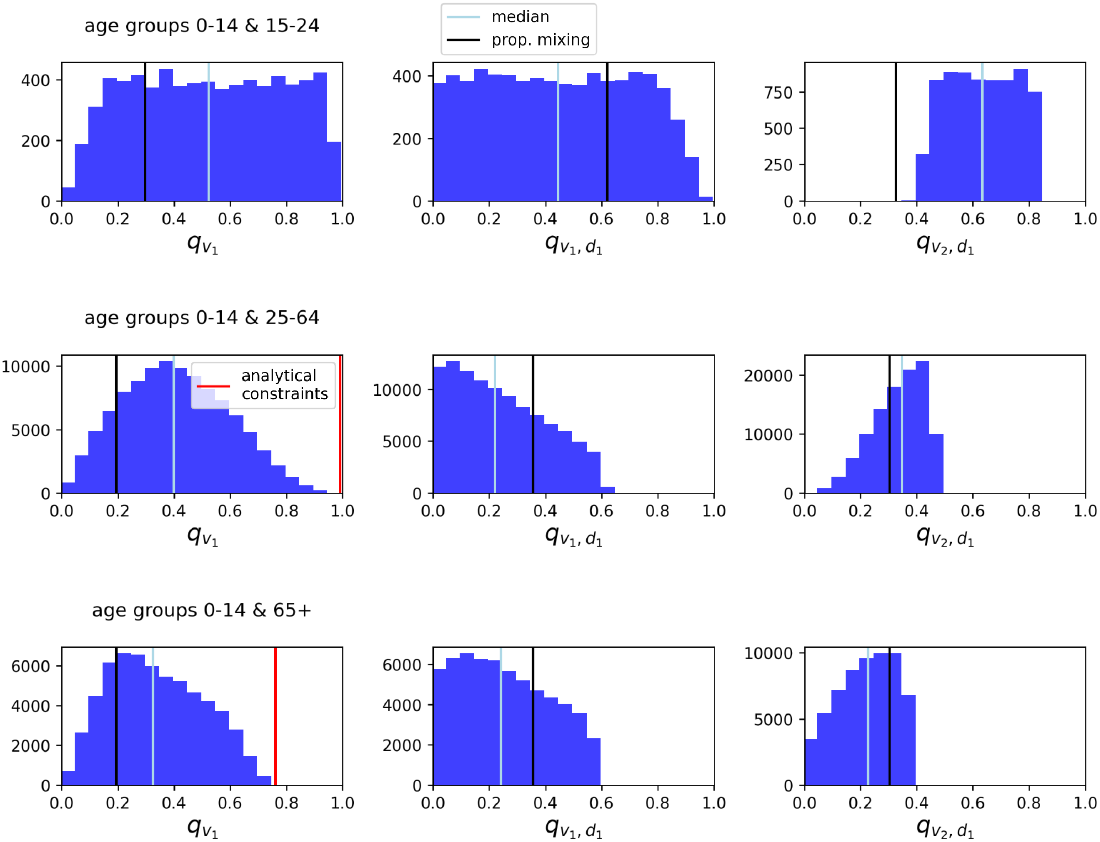
Frequency of values of the 3 free parameters, for each couple of interacting age group (off-diagonal blocks) involving children. In each panel we show the median of the observed distribution (vertical line in light blue), and the value expected from homogeneous mixing, i.e., mixing proportional to the size of the subgroups (vertical black line).

### Assortativity index

In the previous section, we defined assortativity parameters specific for each age group and socio-economic group, required to derive the fully stratified contact matrix. Then we summarised the characteristics of the matrix by defining a single assortativity index (one for education level, and one for SEP), aggregating over age groups and over one of the two social dimensions.

Let us consider the expanded contact matrix 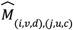.We can aggregate the matrix over the age and education level dimensions, in order to obtain the 2×2 matrix 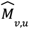, by summing the columns *u, c* and computing a weighted mean on the rows *i, j* as follows:

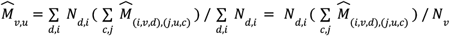

Analogously, we can aggregate the matrix 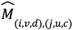 into 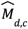 over the age and SEP dimensions.

We then define the assortativity index of the matrix 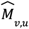 following Ref.^46^ as follows:

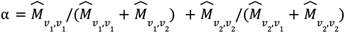

i.e., as the sum of the proportion of contacts on the diagonal, with respect to the total number of contacts per row. If mixing is fully assortative, 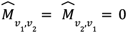 and α = 2; if mixing is completely disassortative, 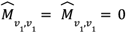 and α = 1; if mixing is homogeneous, i.e., proportional to the size of the groups, then 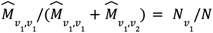,and 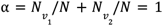

In the main text, we showed the assortativity in the SEP and education level dimensions for each matrix in the set of matrices considered in Fig. 3. In Fig. S13, we show the relative variation in the dominant eigenvalue of the matrix depending on the level of assortativity, computed with respect to a matrix assuming homogeneous mixing. We find that, for both SEP and education level, the dominant eigenvalue is on average slightly higher in the group of matrices with higher assortativity.

**Figure S13.**
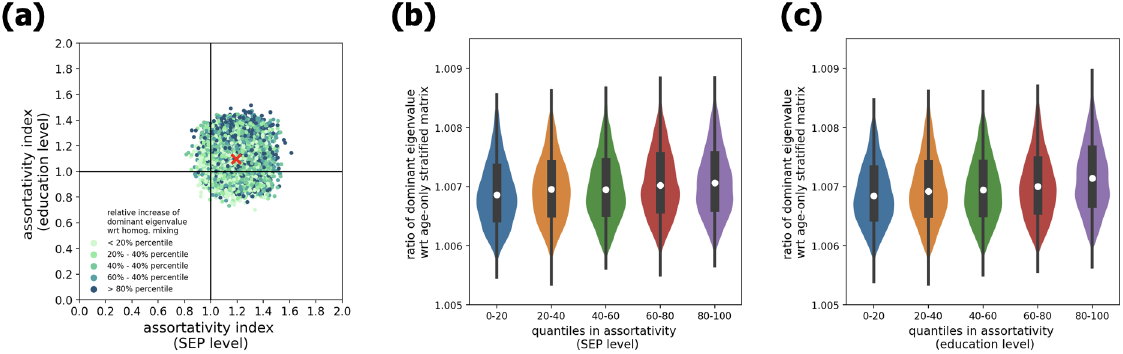
**(a)** Scatter plot of assortativity index in two dimensions (education level and SEP level), color-coded based on ranges of relative increase in the dominant eigenvalue with respect to a homogeneous matrix. **(b-c)** Relative variation in the dominant eigenvalue (with respect to a matrix assuming homogeneous mixing in the SEP and education level dimension) as a function of assortativity in SEP (panel b) and education level (panel c).

### Variation in dominant eigenvalue

In Ref.^34^, it has been shown that (i) 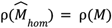,where *M* is the age-stratified matrix, and (ii) 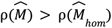 for any matrix 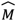 with non-homogeneous mixing on additional dimensions besides age (in our case, SEP and education level). In other words, given the same transmission rate, modeling disease spread with an expanded contact *M* , i.e., accounting for heterogeneous mixing along additional dimensions such as SEP and education level, would result in a higher reproductive number *R* with respect to an age-stratified matrix *M* (or 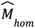 ). On the other hand, given the same *R* , modeling the disease spread assuming the heterogeneous contact matrix 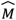 would lead to a lower herd immunity threshold and a lower overall attack rate with respect to using *M* (or 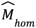), as the additional heterogeneity in the contact patterns along SEP and education would further constrain the age-stratified epidemic^76^.

In Fig. 3f in the main text, we showed the distribution of the ratio of the dominant eigenvalue of the expanded contact matrices with respect to an age-stratified matrix (neglecting the SEP and education level dimension). To quantify the relative role of the two social dimensions, here we show the variation in the dominant eigenvalue in a matrix stratified by age and SEP only, or by age and education level only. We found that introducing the education level dimension, while neglecting the SEP level, would contribute to a higher variation in the dominant eigenvalue with respect to an age-only stratified matrix, with a distribution of the relative increase centred between 0.23 % and 0.36 %. On the other hand, by introducing the stratification by SEP level, neglecting the education level, the distribution of the relative increase in the dominant eigenvalue is wider, ranging between 0.16% and 0.36%, with a median around 0.22%. The results are displayed in Fig. S14.

**Figure S14.**
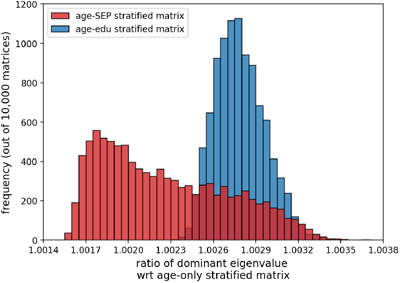
Distribution of the ratio of the dominant eigenvalue of expanded contact matrices that include age and one additional social dimension, with respect to an age-only stratified matrix (i.e., with homogeneous mixing in the SEP and education level dimensions). In red, results for matrices stratified by age and SEP level (neglecting stratification by education level). In blue, results for matrices stratified by age and education level (neglecting stratification by SEP level).

To compare the role of age and socio-economic dimensions, here we show the variation in the dominant eigenvalue in a matrix stratified by age only, by SEP only, by education level only, or by SEP and education level. The variation is computed with respect to a model with no structure (i.e., a constant contact rate for all groups, equal to the overall contact average). The results are displayed in Fig. S15. We found that incorporating the age dimension leads to a variation of 11%, much larger than the variation obtained by including SEP or education level or both socio-economic dimensions. This discrepancy is probably due to two concurrent factors: the fact that assortativity along age is much stronger than assortativity along the socio-economic dimensions, and the fact that the stratification by age introduces more groups (4 age groups) with respect to stratification by SEP and education (two groups).

**Figure S15.**
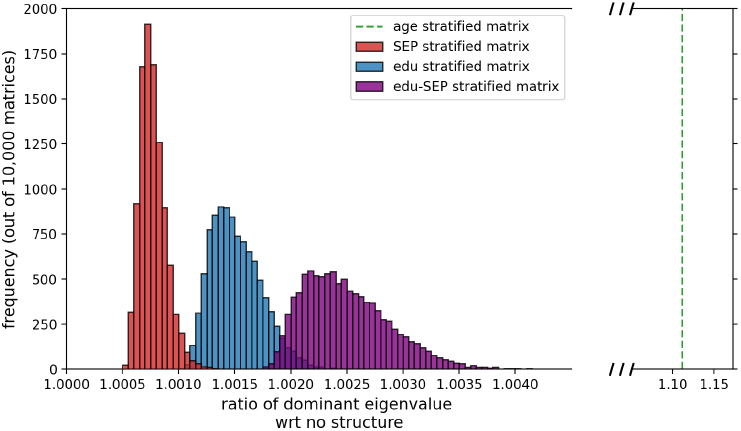
Distribution of the ratio of the dominant eigenvalue of contact matrices, with respect to a model with no contact structure (i.e., a constant contact rate equal to the overall mean number of contacts). In red, results for matrices stratified by SEP level only (size 2×2); in blue, results for matrices stratified by education level only (size 2×2); in purple, results for matrices stratified by SEP and education level (size 4×4); the dashed vertical green line represent the value for the contact matrix stratified by age only (size 4×4).

### Epidemic control

#### Type-reproduction number

For heterogeneous mixing, the type-reproduction number *T*_*g*_ has been introduced as a measure of the control effort needed to contain the epidemic when targeting one specific group *g* of the population^55,56^. Practically, the type-reproduction number can be computed as the dominant eigenvalue of a matrix multiplication involving the next-generation matrix. Control can be implemented either through reduction in susceptibility (S-control, e.g. through vaccination) or through reduction in infectiousness (I-control, e.g. through reduction in contacts or shortening of the infectious period).

Assuming an index case of type 1, the type-reproduction number *T*_1_ denotes the cumulative number of infected hosts of type 1 resulting from all chains of infection, without another infected host of type 1 being allowed to reproduce. The type-reproduction number *T*_1_ for one particular host group can be generalized to *T*_*g*_ to provide insights on the control effort needed to halt disease spread when targeting a subset of *g* host types. The epidemic is controlled if a proportion of group *l* greater than 1 − 1/*T*_*g*_ is permanently immune or fully isolated at the start of the epidemic. Thus, the quantity 1 − 1/*T*_*g*_ corresponds to the immunity threshold specific to some target group(s).

#### Epidemic scenario with homogenous susceptibility

In this section, we report the analysis of the effectiveness of targeted control strategies in an epidemic scenario with homogeneous susceptibility (Fig. S16), as done in the main text with the scenario with 50% reduction in susceptibility for children (Fig. 5).

In a scenario with homogeneous susceptibility, we found that children aged 0-14 play a major role in transmission (around ∼50% contribution to *R*_0_ accounting for both low SEP and high SEP), as shown in Fig. 4 in the main text. Therefore, on one hand, we would expect that a targeted strategy that includes children would be highly effective, and on the other hand, excluding children would hardly result in a successful control strategy. This is indeed what we found in Fig. S16. We found that a strategy targeting individuals with middle low education is effective for 100% of the contact matrices considered (Fig. S16i) and requires a low control effort (median 30%, 95% probability range 29% - 33%, Fig. S11d). Instead, a strategy targeted at individuals with a high education level (hence excluding all children) is never effective in preventing disease spread, regardless of the control effort (Fig. S16f,i). For a strategy targeted at individuals with high SEP and middle-low education level, we found that a gradient in the control effort required as a function of the assortativity index in the education level and SEP dimensions (Fig. S16h), in line with what we found in the main text (Fig. 5h). The lower the assortativity in contacts in the additional social dimensions, the higher the chances that the strategy would be successful and the lower the control effort required to control the epidemic.

**Figure S16.**
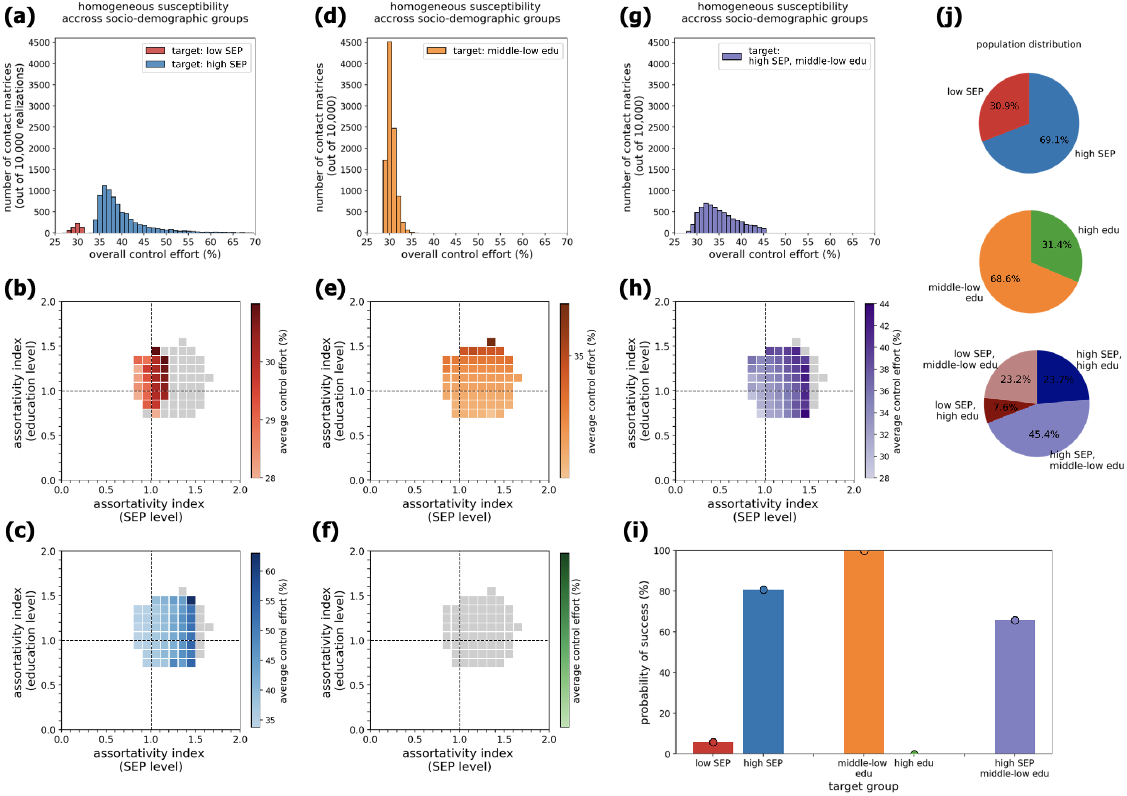
Effective targeted control strategies. We considered a structured SIR epidemic model, stratified by age and socio-economic levels with heterogeneous mixing, and homogeneous susceptibility across age and socio-economic groups. We assumed *R*_0_ = 1. 5, and an average infectious period of 3 days. **(a)** Distribution of the overall control effort required by strategies targeted at individuals with low SEP (red) or with high SEP (blue). **(b)** Assortativity levels in the SEP and education dimensions for the subset of matrices which allow effective control (colored cells) and those matrices for which the strategy would not be effective (grey cells). The color gradient indicates the average control effort required for a given range of assortativity. The strategy considered here is targeted at the low SEP group. **(c)** As in panel (b), but considering a strategy targeted at the group with high SEP. **(c)** Distribution of the overall control effort required by strategies targeted at individuals with middle-low (orange) or with high education level (green). **(e)** As in panel (b), but considering a strategy targeted at the group with middle-low education. **(f)** As in panel (b), but considering a strategy targeted at the group with high education. **(g)** Distribution of the overall control effort required by strategies targeted at individuals with high SEP and middle-low education level. **(h)** As in panel (b), but considering a strategy targeted at the group with high SEP and middle-low education level. **(i)** Probability of success of the targeted strategy, defined as the fraction of contact matrices for which there exists a critical control effort which allows epidemic control. **(j)** Pie charts displaying the distribution of the population in three partitions, i.e., low SEP/high SEP (top), middle-low education/high education (center), and the combination of the two dimensions.

## References

1. Jit, M. et al. Reflections On Epidemiological Modeling To Inform Policy During The COVID-19 Pandemic In Western Europe, 2020–23. Health Aff. (Millwood) 42, 1630–1636 (2023).

2. Willem, L. et al. The impact of quality-adjusted life years on evaluating COVID-19 mitigation strategies: lessons from age-specific vaccination roll-out and variants of concern in Belgium (2020-2022). BMC Public Health 24, 1171 (2024).

3. Klinkenberg, D., Backer, J., De Keizer, N. & Wallinga, J. Projecting COVID-19 intensive care admissions for policy advice, the Netherlands, February 2020 to January 2021. Eurosurveillance 29, (2024).

4. Pullano, G. et al. Underdetection of cases of COVID-19 in France threatens epidemic control. Nature 590, 134–139 (2021).

5. Davies, N. G. et al. Association of tiered restrictions and a second lockdown with COVID-19 deaths and hospital admissions in England: a modelling study. Lancet Infect. Dis. 0, (2020).

6. Zhang, J. et al. Changes in contact patterns shape the dynamics of the COVID-19 outbreak in China. Science 368, 1481–1486 (2020).

7. Verelst, F. et al. SOCRATES-CoMix: a platform for timely and open-source contact mixing data during and in between COVID-19 surges and interventions in over 20 European countries. BMC Med. 19, 254 (2021).

8. Di Domenico, L., Bosetti, P., Sabbatini, C. E., Opatowski, L. & Colizza, V. Mobility-driven synthetic contact matrices: a scalable solution for real-time pandemic response modeling. 2024.12.12.24318903 Preprint at 10.1101/2024.12.12.24318903 (2024).

9. Manna, A., Koltai, J. & Karsai, M. Importance of social inequalities to contact patterns, vaccine uptake, and epidemic dynamics. Nat. Commun. 15, 4137 (2024).

10. Backer, J. A. et al. Contact behaviour before, during and after the COVID-19 pandemic in the Netherlands: evidence from contact surveys, 2016 to 2017 and 2020 to 2023. Eurosurveillance 29, (2024).

11. Drolet, M. et al. Time trends in social contacts before and during the COVID-19 pandemic: the CONNECT study. BMC Public Health 22, 1032 (2022).

12. Wambua, J. et al. The influence of COVID-19 risk perception and vaccination status on the number of social contacts across Europe: insights from the CoMix study. BMC Public Health 23, 1350 (2023).

13. Jarvis, C. I. et al. Social contact patterns following the COVID-19 pandemic: a snapshot of post-pandemic behaviour from the CoMix study. Epidemics 48, 100778 (2024).

14. Wong, K. L. M. et al. Social contact patterns during the COVID-19 pandemic in 21 European countries – evidence from a two-year study. BMC Infect. Dis. 23, 268 (2023).

15. Godbout, A. et al. Time trends in social contacts of individuals according to comorbidity and vaccination status, before and during the COVID-19 pandemic. BMC Med. 20, 199 (2022).

16. Quaife, M. et al. COVID-19 vaccine hesitancy and social contact patterns in Pakistan: results from a national cross-sectional survey. BMC Infect. Dis. 23, 321 (2023).

17. Valdano, E., Lee, J., Bansal, S., Rubrichi, S. & Colizza, V. Highlighting socio-economic constraints on mobility reductions during COVID-19 restrictions in France can inform effective and equitable pandemic response. J. Travel Med. 28, (2021).

18. Heroy, S., Loaiza, I., Pentland, A. & O’Clery, N. COVID-19 policy analysis: labour structure dictates lockdown mobility behaviour. J. R. Soc. Interface 18, 20201035 (2021).

19. Jay, J. et al. Neighbourhood income and physical distancing during the COVID-19 pandemic in the United States. Nat. Hum. Behav. 4, 1294–1302 (2020).

20. Riou, J. et al. Socioeconomic position and the COVID-19 care cascade from testing to mortality in Switzerland: a population-based analysis. Lancet Public Health 6, e683–e691 (2021).

21. Sharma, S. et al. Sociodemographic differences in the response to changes in COVID-19 testing guidelines. Eur. J. Public Health ckae145 (2024) doi:10.1093/eurpub/ckae145.

22. Nordberg, P. et al. Immigrant background and socioeconomic status are associated with severe COVID-19 requiring intensive care. Sci. Rep. 12, 12133 (2022).

23. Reichmuth, M. L. et al. Socio-demographic characteristics associated with COVID-19 vaccination uptake in Switzerland: longitudinal analysis of the CoMix study. BMC Public Health 23, 1523 (2023).

24. Ligeti, A. S. et al. Socioeconomic determinants and reasons for non-acceptance to vaccination recommendations during the 3rd - 5th waves of the COVID-19 pandemic in Hungary. BMC Public Health 24, 1796 (2024).

25. Barceló, M. A., Perafita, X. & Saez, M. Spatiotemporal variability in socioeconomic inequalities in COVID-19 vaccination in Catalonia, Spain. Public Health 227, 9–15 (2024).

26. Charland, K. et al. Parental decisions regarding the vaccination of children and adolescents against SARS-CoV-2 from 2020 to 2023: A descriptive longitudinal study of parents and children in Montreal, Canada. Vaccine 43, 126489 (2025).

27. Sacre, A. et al. Socioeconomic inequalities in vaccine uptake: A global umbrella review. PLOS ONE 18, e0294688 (2023).

28. Ayorinde, A. et al. Health inequalities in infectious diseases: a systematic overview of reviews. BMJ Open 13, e067429 (2023).

29. Tizzoni, M. et al. Addressing the socioeconomic divide in computational modeling for infectious diseases. Nat. Commun. 13, 2897 (2022).

30. Naidoo, M. et al. Incorporating social vulnerability in infectious disease mathematical modelling: a scoping review. BMC Med. 22, 125 (2024).

31. Zelner, J. et al. There are no equal opportunity infectors: Epidemiological modelers must rethink our approach to inequality in infection risk. PLOS Comput. Biol. 18, e1009795 (2022).

32. Buckee, C., Noor, A. & Sattenspiel, L. Thinking clearly about social aspects of infectious disease transmission. Nature 595, 205–213 (2021).

33. Bedson, J. et al. A review and agenda for integrated disease models including social and behavioural factors. Nat. Hum. Behav. 5, 834–846 (2021).

34. Manna, A., Dall’Amico, L., Tizzoni, M., Karsai, M. & Perra, N. Generalized contact matrices for epidemic modeling. Sci. Adv. 10, (2024).

35. Manna, A., Karsai, M. & Perra, N. Social inequalities in vaccine coverage and their effects on epidemic spreading. 2024.11.01.24316556 Preprint at 10.1101/2024.11.01.24316556 (2024).

36. Panczak, R. et al. A Swiss neighbourhood index of socioeconomic position: development and association with mortality. J. Epidemiol. Community Health 66, 1129–1136 (2012).

37. Panczak, R., Berlin, C., Voorpostel, M., Zwahlen, M. & Egger, M. The Swiss neighbourhood index of socioeconomic position: update and re-validation. Swiss Med. Wkly. 153, 40028–40028 (2023).

38. Riesen, M. et al. Exploring variation in human papillomavirus vaccination uptake in Switzerland: a multilevel spatial analysis of a national vaccination coverage survey. BMJ Open 8, e021006 (2018).

39. Reichmuth, M. L. et al. Social contacts in Switzerland during the COVID-19 pandemic: insights from the CoMix study. Epidemics 100771 (2024) doi:10.1016/j.epidem.2024.100771.

40. Reichmuth, M. L., Heron, L., Low, N. & Althaus, C. L. CoMix social contact data for Switzerland (2021-2022). Zenodo 10.5281/zenodo.10147647 (2023).

41. State Secretariat for Education, Research and Innovation (SERI). Swiss Education System. https://www.sbfi.admin.ch/sbfi/en/home/bildung/bildungsraum-schweiz/das-duale-system.html.

42. Federal Statistical Office. Population. https://www.bfs.admin.ch/bfs/en/home/statistiken/bevoelkerung.html.

43. Federal Statistical Office. Permanent resident population by age, canton, district and commune, 2010-2022. https://www.bfs.admin.ch/asset/en/26565304.

44. Federal Statistical Office. Highest completed education by various socio-demographic characteristics in Switzerland, 2010-2022. https://www.bfs.admin.ch/asset/en/30148622.

45. Mossong, J. et al. Social contacts and mixing patterns relevant to the spread of infectious diseases. PLoS Med. 5, e74 (2008).

46. Garnett & Anderson. Factors controlling the spread of HIV in heterosexual communities in developing countries: patterns of mixing between different age and sexual activity classes. Philos. Trans. R. Soc. Lond. B. Biol. Sci. 342, 137–159 (1993).

47. Valle, S. Y. D., Hyman, J. M. & Chitnis, N. Mathematical models of contact patterns between age groups for predicting the spread of infectious diseases. Math. Biosci. Eng. 10, 1475–1497 (2013).

48. Kermack, W. O. & McKendrick, A. G. A contribution to the mathematical theory of epidemics. Proc. R. Soc. Lond. 115, 700–721 (1927).

49. Wallinga, J., Teunis, P. & Kretzschmar, M. Using Data on Social Contacts to Estimate Age-specific Transmission Parameters for Respiratory-spread Infectious Agents. Am. J. Epidemiol. 164, 936–944 (2006).

50. Diekmann, O., Heesterbeek, J. A. P. & Metz, J. A. J. On the definition and the computation of the basic reproduction ratio R0 in models for infectious diseases in heterogeneous populations. J. Math. Biol. 28, 365–382 (1990).

51. van den Driessche, P. & Watmough, J. Reproduction numbers and sub-threshold endemic equilibria for compartmental models of disease transmission. Math. Biosci. 180, 29–48 (2002).

52. Diekmann, O., Heesterbeek, J. A. P. & Roberts, M. G. The construction of next-generation matrices for compartmental epidemic models. J. R. Soc. Interface 7, 873–885 (2010).

53. Angeli, L. et al. Who acquires infection from whom? A sensitivity analysis of transmission dynamics during the early phase of the COVID-19 pandemic in Belgium. J. Theor. Biol. 581, 111721 (2024).

54. Hethcote, H. W. The Mathematics of Infectious Diseases. SIAM Rev. 42, 599–653 (2000).

55. Roberts, M. G. & Heesterbeek, J. A. P. A new method for estimating the effort required to control an infectious disease. Proc. R. Soc. Lond. B Biol. Sci. 270, 1359–1364 (2003).

56. Heesterbeek, J. A. P. & Roberts, M. G. The type-reproduction number T in models for infectious disease control. Math. Biosci. 206, 3–10 (2007).

57. Munday, J. D. et al. Estimating the impact of reopening schools on the reproduction number of SARS-CoV-2 in England, using weekly contact survey data. BMC Med. 19, 233 (2021).

58. Quaife, M. et al. The impact of COVID-19 control measures on social contacts and transmission in Kenyan informal settlements. BMC Med. 18, 316 (2020).

59. Duran-Sala, M. et al. Disentangling individual-level from location-based income uncovers socioeconomic preferential mobility and impacts segregation estimates. Preprint at 10.48550/arXiv.2407.01799 (2024).

60. Goodfellow, L., van Leeuwen, E. & Eggo, R. M. COVID-19 inequalities in England: a mathematical modelling study of transmission risk and clinical vulnerability by socioeconomic status. BMC Med. 22, 162 (2024).

61. Moro, E., Calacci, D., Dong, X. & Pentland, A. Mobility patterns are associated with experienced income segregation in large US cities. Nat. Commun. 12, 4633 (2021).

62. Munday, J. D., van Hoek, A. J., Edmunds, W. J. & Atkins, K. E. Quantifying the impact of social groups and vaccination on inequalities in infectious diseases using a mathematical model. BMC Med. 16, 162 (2018).

63. Zipfel, C. M., Colizza, V. & Bansal, S. Health inequities in influenza transmission and surveillance. PLOS Comput. Biol. 17, e1008642 (2021).

64. Britton, T. & Ball, F. Improving the use of social contact studies in epidemic modelling. Preprint at 10.48550/arXiv.2408.07298 (2024).

65. Kiss, I. Z., Green, D. M. & Kao, R. R. The effect of network mixing patterns on epidemic dynamics and the efficacy of disease contact tracing. J. R. Soc. Interface 5, 791–799 (2008).

66. Nishiura, H., Cook, A. R. & Cowling, B. J. Assortativity and the Probability of Epidemic Extinction: A Case Study of Pandemic Influenza A (H1N1-2009). Interdiscip. Perspect. Infect. Dis. 2011, 194507 (2011).

67. Geismar, C., White, P. J., Cori, A. & Jombart, T. Sorting out assortativity: when can we assess the contributions of different population groups to epidemic transmission? 2024.03.13.24304225 Preprint at 10.1101/2024.03.13.24304225 (2024).

68. Ma, K. C., Menkir, T. F., Kissler, S., Grad, Y. H. & Lipsitch, M. Modeling the impact of racial and ethnic disparities on COVID-19 epidemic dynamics. eLife 10, e66601 (2021).

69. Prem, K. et al. Projecting contact matrices in 177 geographical regions: An update and comparison with empirical data for the COVID-19 era. PLOS Comput. Biol. 17, e1009098 (2021).

70. Panczak, R., Berlin, C., Voorpostel, M., Zwahlen, M. & Egger, M. Product: Swiss Neighbourhood Index of Socioeconomic Position (Swiss-SEP). https://boris-portal.unibe.ch/entities/product/8fb9dd20-d609-4b09-950b-ae74d242544a (2022).

71. Federal Statistical Office. Population and household statistics (STATPOP), geodata 2022. https://www.bfs.admin.ch/asset/en/27965868.

72. Hale, T. et al. A global panel database of pandemic policies (Oxford COVID-19 Government Response Tracker). Nat. Hum. Behav. 5, 529–538 (2021).

73. Google. COVID-19 Community Mobility Report. COVID-19 Community Mobility Report https://www.google.com/covid19/mobility?hl=en.

74. Federal Statistical Office. ThemaKart map boundaries - Set 2023. https://www.bfs.admin.ch/asset/en/24025646.

75. Federal Office of Topography (swisstopo). Population (residents). https://map.geo.admin.ch/#/map?lang=en&center=2533675.22,1170896.93&z=2.479&topic=ech&layers=ch.swisstopo.zeitreihen@year=1864,f;ch.bfs.gebaeude_wohnungs_register,f;ch.bav.haltestellen-oev,f;ch.swisstopo.swisstlm3d-wanderwege,f;ch.bfs.volkszaehlung-bevoelkerungsstatistik_einwohner@year=2021;ch.bak.schutzgebiete-unesco_weltkulturerbe,f&bgLayer=ch.swisstopo.pixelkarte-farbe.

76. Britton, T., Ball, F. & Trapman, P. A mathematical model reveals the influence of population heterogeneity on herd immunity to SARS-CoV-2. Science 369, 846–849 (2020).

